# Comparative efficacy and acceptability of cognitive-behavioural therapy for insomnia and its abbreviated versions: a systematic review and network meta-analysis

**DOI:** 10.64898/2026.07.04.26357278

**Authors:** Masatsugu Sakata, Shino Kikuchi, Masami Ito, Rie Toyomoto, Hikari N. Takashina, Shintaro Hara, Ryuichiro Yamamoto, Shun Nakajima, Hiroku Noma, Kota Imai, Shunichi Sato, Daiki Nagaoka, Yusuke Takahashi, Keita Kawai, Seina Shinno, Azusa Ishii, Michael Perlis, Cagdas Türkmen, Elisabeth Hertenstein, Annemieke van Straten, Yuki Furukawa, the SLEEPI investigators

## Abstract

**Objective:** To assess the comparative efficacy and acceptability of cognitive behavioural therapy for insomnia (CBT-I), its abbreviated versions and control conditions.

**Design:** Systematic review and network meta-analysis.

**Methods:** Screening, data extraction, coding, and risk of bias assessment were performed independently and in duplicate. Frequentist, random-effects network meta-analyses estimated odds ratios (ORs) or mean differences with 95% confidence intervals (CIs). The primary outcome was insomnia remission post-treatment. Secondary outcomes included dropout and subjective sleep continuity measures. Quality of the evidence for each arm was graded using the confidence in network meta-analysis (CINeMA).

**Data sources:** We searched MEDLINE, Embase, PsycINFO and Cochrane CENTRAL from inception to December 15, 2025, with a medical information specialist.

**Eligibility criteria for selecting studies:** Randomized –controlled trials (RCTs) comparing CBT-I and its abbreviated versions with each other or with control conditions, in adults with insomnia, with or without comorbidities. To reduce clinical heterogeneity related to treatment intensity and adherence, we restricted inclusion to in-person delivery.

**Results:** We identified 11,379 records and included 77 RCTs (5,731 participants; mean age 52.2 years; 3,473 female). CBT-I (number of arms k = 53; number of participants n = 2,002), sleep restriction and stimulus control therapy (SRT&SCT; k = 16; n = 549), sleep restriction therapy (SRT; k = 5; n = 196) and stimulus control therapy (SCT; k = 7; n = 144) were associated with higher remission than sleep hygiene, relaxation therapy and other control conditions. These interventions were also effective in improving subjective sleep continuity measures. Cognitive therapy for insomnia (CT-I) was more beneficial than relaxation therapy. Dropout did not differ meaningfully between interventions and controls. Confidence in evidence was moderate for CBT-I, low for SRT&SCT and SRT, very low for SCT. Given the weighted mean proportion of insomnia remission among sleep hygiene arms of 20%, CBT-I probably leads to a remission rate of 41% (95% CI, 34%; 48%), SRT&SCT may lead to a remission rate of 40% (30%; 52%), SCT 43% (25%; 63%), and SRT 41% (26%; 57%).

**Conclusions:** CBT-I doubles the absolute insomnia remission compared with sleep hygiene, and its abbreviated behavioural therapies, namely, SRT&SCT, SCT and SRT may offer similar benefits with lower resource requirements, but evidence is less certain. CT-I needs further investigations. Relaxation therapy was inferior to these therapies. Implementation decisions should consider resource requirements and evidence certainty.

**What is already known on this topic:** - Insomnia is prevalent and disabling, and cognitive behavioural therapy for insomnia (CBT-I) is recommended as the first-line treatment.

- CBT-I and its abbreviated versions are recommended in guidelines, but their comparative efficacy and acceptability remain uncertain.

**What this study adds:** - CBT-I and its core behavioural components (sleep restriction and stimulus control) probably achieve similar remission rates, offering scalable options where full CBT-I is not available.

- Relaxation therapy was inferior to cognitive behavioural therapy for insomnia, its abbreviated, behavioural interventions, and cognitive therapy for insomnia.

- Dissemination and implementation attempts should balance the confidence in the evidence and simplicity of abbreviated versions.

## INTRODUCTION

Insomnia affects around 12% of the adult population,[1] and is associated with daytime impairment.[2] It is also associated with an increased risk of various physical and mental disorders.[3] Cognitive-behavioural therapy for insomnia (CBT-I) is recommended as the first-line treatment and is more effective than pharmacotherapy.[4–6] However, access to CBT-I remains limited in most of the clinical settings, and medications continue to be widely used despite concerns about adverse events,[7,8] and uncertain long-term benefits.[9]

To address limitations in availability, simplified behavioural interventions, here referred to as abbreviated behavioural interventions, which include sleep restriction therapy (SRT) and/or stimulus control therapy (SCT) without the full multicomponent CBT-I package, have been proposed as resource-saving alternatives and acknowledged in guidelines,[10,11] Table 1 summarises the characteristics of CBT-I components.[12] Most contemporary guidelines endorse multicomponent CBT-I as the preferred package. However, several guidelines, including the American Academy of Sleep Medicine and the European Insomnia Guideline, also acknowledge abbreviated behavioural interventions such as SCT and SRT as acceptable alternatives under certain circumstances. [10,11] This variation reflects ongoing uncertainty regarding the extent to which full multicomponent package is necessary to achieve remission.

**Table 1.** Summary of interventions.

| Intervention | Description |
| --- | --- |
| Educational components |  |

| **Intervention** | **Description** |
| --- | --- |
| **Sleep hygiene education** | General explanation about sleep (e.g., sleep biology, characteristics of healthy sleep, changes in sleep patterns with aging) and general recommendations about lifestyle (e.g., exercise, diet, substance use) and environmental factors (e.g., bed partner, light, noise, temperature) to improve sleep. This may include some elements of other components, e.g. stimulus control, but those should not be the predominant part of the intervention. |
| **Cognitive components** |  |
| **Cognitive restructuring** | Skills to identify, challenge and modify unhelpful beliefs about sleep that may disturb sleep. Sometimes simply called cognitive therapy. |
| **Behavioural components** |  |
| **Sleep restriction** | Skills to improve sleep by limiting time in bed. First, time in bed is restricted to the average subjective sleep duration and then it is increased or decreased depending on sleep efficiency. |
| **Stimulus control** | Skills to re-associate the bed with sleep. Patients are instructed to get up at the same time every morning, refrain from daytime napping, go to bed only when sleepy, get out of bed when unable to sleep, and use the bed/bedroom for sleep and sex only. |
| **Relaxation** | Structured exercises to reduce somatic tension (e.g., abdominal breathing, progressive muscle relaxation, autogenic training) and cognitive arousal (e.g., guided imagery training). |
| **Others** |  |
| **Waiting component** | Participants are aware that they can receive an active treatment after a waiting phase. This condition is known to have a nocebo effect. |

Behavioural treatment for insomnia has evolved over several decades. In the 1970s and 80s, SCT and SRT were introduced as stand-alone interventions.[13,14] These approaches were later integrated with cognitive techniques into multicomponent CBT-I, which became the standard format endorsed by guidelines.[15,16] More recently, workforce constraints and service pressures have renewed interest in stand-alone, or simplified behavioural formats as potentially scalable first-line options.[17,18] Contemporary evidence supports the efficacy of sleep restriction and stimulus control delivered individually, and pragmatic trials suggest that brief behavioural interventions can be implemented in primary care settings. [19] Component network meta-analyses showed that sleep restriction, stimulus control, and cognitive restructuring represent key components of CBT-I.[12,20] Although multicomponent CBT-I is effective, implementing the full treatment package in routine clinical practice can be challenging because it requires multiple components and substantial therapist time. Identifying whether simpler behavioural packages achieve comparable remission may therefore help inform scalable treatment delivery. However, these analyses rely on the statistical assumption that treatment effects are additive and that interactions between the components are minimal. [12,20] Although sensitivity analyses have explored potential interactions, this does not guarantee the non-existence of meaningful interactions.[12]

Therefore, we conducted a network meta-analysis comparing CBT-I and its abbreviated versions, using remission as the primary outcome, with the aim of informing decisions about scalable treatment delivery in routine care.

## METHODS

We followed the Preferred Reporting Items for Systematic reviews and Meta-Analyses (PRISMA) guideline extension for NMA.[21] This project was part of the Systematic Living Evidence Evaluation of Psychotherapies for Insomnia (SLEEPI; https://osf.io/c82xu/). The protocol of this study was prospectively registered in the Open Science Framework (https://osf.io/z48r2/). People with lived experience were not involved in the research process.

### Eligibility criteria and search strategies

#### Study design and publication type

We included randomised controlled trials that compared CBT-I to its abbreviated version or a control condition in the treatment of adults with chronic insomnia. We restricted inclusion to randomised controlled trials to obtain the most reliable estimates of intervention. We focused on peer-reviewed English-language studies to ensure methodological quality and feasibility of accurate assessment. Conference abstracts, dissertations, preprints and results posted only on registries were not included. These decisions were made in consultation with the METAPSY project (https://www.metapsy.org/), [22] which is building living systematic reviews covering a broad range of psychotherapies across an entire research domain.

#### Participants

We included trials on adults with insomnia either diagnosed according to the formal diagnostic criteria (such as the Diagnostic and Statistical Manual of Mental Disorders, the International Classification of Diseases or the International Classification of Sleep Disorders) or judged so by elevated symptoms scales (e.g., Insomnia Severity Index). The criteria needed to include daytime impairment. We included patients with psychiatric or physical comorbidities.[23]

#### Interventions and controls

We defined CBT-I as interventions including sleep restriction, stimulus control and cognitive restructuring. Additional elements (e.g., relaxation) were permitted. In this study, we focused on conventional CBT-I and excluded third-wave adaptations such as mindfulness-based or acceptance and commitment therapy.

We classified abbreviated formats according to their behavioral content: combined sleep restriction therapy and stimulus control therapy (SRT&SCT); sleep restriction therapy alone (SRT); stimulus control therapy alone (SCT); cognitive therapy alone (CT-I); and relaxation therapy (RT). Control conditions included waiting list, sleep hygiene/psychoeducation, attention/psychological placebo control, or no treatment.

To enhance comparability, we included only in-person intervention, as delivery mode may influence efficacy.[12,24] Concomitant pharmacotherapies were permitted if balanced across arms. We included interventions of any duration.

Where trials included multiple eligible arms, we included only the relevant comparisons and combined arms sharing identical components.

### Primary outcome and secondary outcomes

The primary outcome of interest in this study is treatment efficacy at post treatment or at its closest time point. We used the remission, defined as reaching a satisfactory state at endpoint measured by any validated self-reported scale (dichotomous). Secondary outcomes included treatment acceptability, assessed by all-cause dropout rates (dichotomous); sleep diary-derived measures, including sleep efficiency (SE, %), sleep latency (SL, minutes), wake after sleep onset (WASO, minutes), and total sleep time (TST, minutes); and treatment efficacy at long-term follow-up, defined as the longest follow-up between 3 and 12 months (dichotomous).

We prioritized the end score over the change score,[25] and methods accounting for missing outcome data (e.g., mixed-models of repeated measurement, multiple imputations) over last observation carried forward and over observed cases. We used the number of participants randomised as the denominator for dichotomous outcomes. We used odds ratio for dichotomous outcomes, mean difference for continuous outcomes expressed in minutes and percent.

### Search methods for identification of studies

We used our SLEEPI database, which is based on regular updates of comprehensive literature search in MEDLINE, Embase, PsycINFO and Cochrane CENTRAL from inception until December 15, 2025, in collaboration with a medical information specialist. The following terms were used (including synonyms and closely related words) as index terms or free-text words: “Insomnia”, “Sleep disturbance”, “Cognitive behavioral therapy”, “Clinical trials”.

The references of the identified articles were searched for relevant publications. Duplicate articles were excluded by a medical information specialist using Endnote X21.0.1 (Clarivate), following the Amsterdam Efficient Deduplication (AED)-method[26] and the Bramer-method[27].

More details can be found in eAppendix and the protocol of the SLEEPI project in the Open Science Framework (https://osf.io/c82xu/).

### Selection process and data extraction

Two reviewers (pairs of DN, HN, HNT, KI, KK, MI, MS, RT, RY, SH, SK, SN, SS, YF, or YT) independently screened all potentially relevant titles and abstracts for eligibility. The full texts of the selected articles were obtained for further full text review by two independent reviewers (pairs of AI, HN, HNT, KK, MI, MS, RT, RY, SH, SK, SN, SS, YF, or YT). Two independent reviewers (pairs of MI, MS, SK, SH, RT, or RY) extracted relevant data, and evaluated the risk of bias of individual studies using the revised Cochrane Risk of Bias assessment tool (RoB2) for the primary outcome. [28] Differences in judgements were resolved through discussion.

### Data analysis

We created a network diagram to see the overall geometry of the network. We then examined the transitivity assumption by creating a table of important trial and patient characteristics to see if potential effect modifiers (age, sex, baseline severity) were similarly distributed among comparisons. We imputed missing standard deviations from other trials,[29] and imputed insomnia remission using validated methods.[12,30] Remission was defined according to the thresholds specified for each validated instrument used in the original trials. Given the expected clinical and methodological heterogeneity of treatment effects among the studies, we used the random-effects model. The variance structure was specified using a common between-study heterogeneity variance across the network, assuming a common τ² for all comparisons. We checked the consistency of the network using local[31] and global[32] inconsistency tests. We visualised the NMA results using psychoeducation (sleep hygiene education) as the reference, and ordering interventions according to the P score [33]. We summarised the primary and secondary outcomes using the Kilim plot [34] We performed several sensitivity analyses to confirm the robustness of the primary analysis: excluding trials without formal diagnosis of insomnia; excluding trials focusing on patients with comorbidities; excluding trials with 20% or higher dropout rate; excluding trials with high overall risk of bias; and focusing on active intervention arms (CBT-I, SRT&SCT, SRT, SCT, CT-I and RT).

We assessed the presence of small study effects using the contour-enhanced funnel plot, including publication bias, for comparisons with more than 10 trials. We assessed the certainty of evidence in network estimates of the primary outcome using the Confidence in Network Meta-Analysis (CINeMA) framework.[35]

Using the control insomnia remission rate among psychoeducation arms and the result of NMA, we calculated the insomnia remission rates among other interventions to increase the interpretability.[36]

We performed the analysis in *R* (version 4.5.1, R foundation, Vienna, Austria) [37] using the *netmeta* (version 3.2-0) package [38] and the *meta* (version 8.2-1) package [39].

### Patient and public involvement

There was no patient or public involvement in the development of this manuscript.

## RESULTS

We identified 11,379 references and screened 6,851 references after removing duplicates. Figure 1 shows the PRISMA flow diagram. We included 77 trials with 5,731 participants. eAppendix2 presents the list of included trials. Typical participants were females in their forties and fifties (3473 females, 2258 males; mean age, 52.2 years [standard deviation (SD), 15.5]; Ethnicity data not available). We included 53 CBT-I arms (number of allocated participants = 2,002), 16 SRT&SCT arms (n = 549), 5 SRT arms (n = 196), 7 SCT arms (n = 144), 2 CT-I arms (n = 108), and 9 relaxation therapy arms (n = 270). Formal diagnostic criteria were used in 44 trials and elevated insomnia rating scales were used in 45 trials. Comorbidities were allowed in 62 trials and excluded in 15 trials. Co-administration of hypnotics were allowed in 50 trials and not allowed in 27 trials. The overall risk of bias for the primary outcome was low in 12 trials, some concerns in 35 trials, and high in 30 trials. eAppendix2 shows the study and patient characteristics and the RoB2 evaluation. The network for the primary outcome was well connected (Figure 2). There was no clear evidence of unequal distribution of effect modifiers across the comparisons. (eAppendix 3)

**Figure 1.**
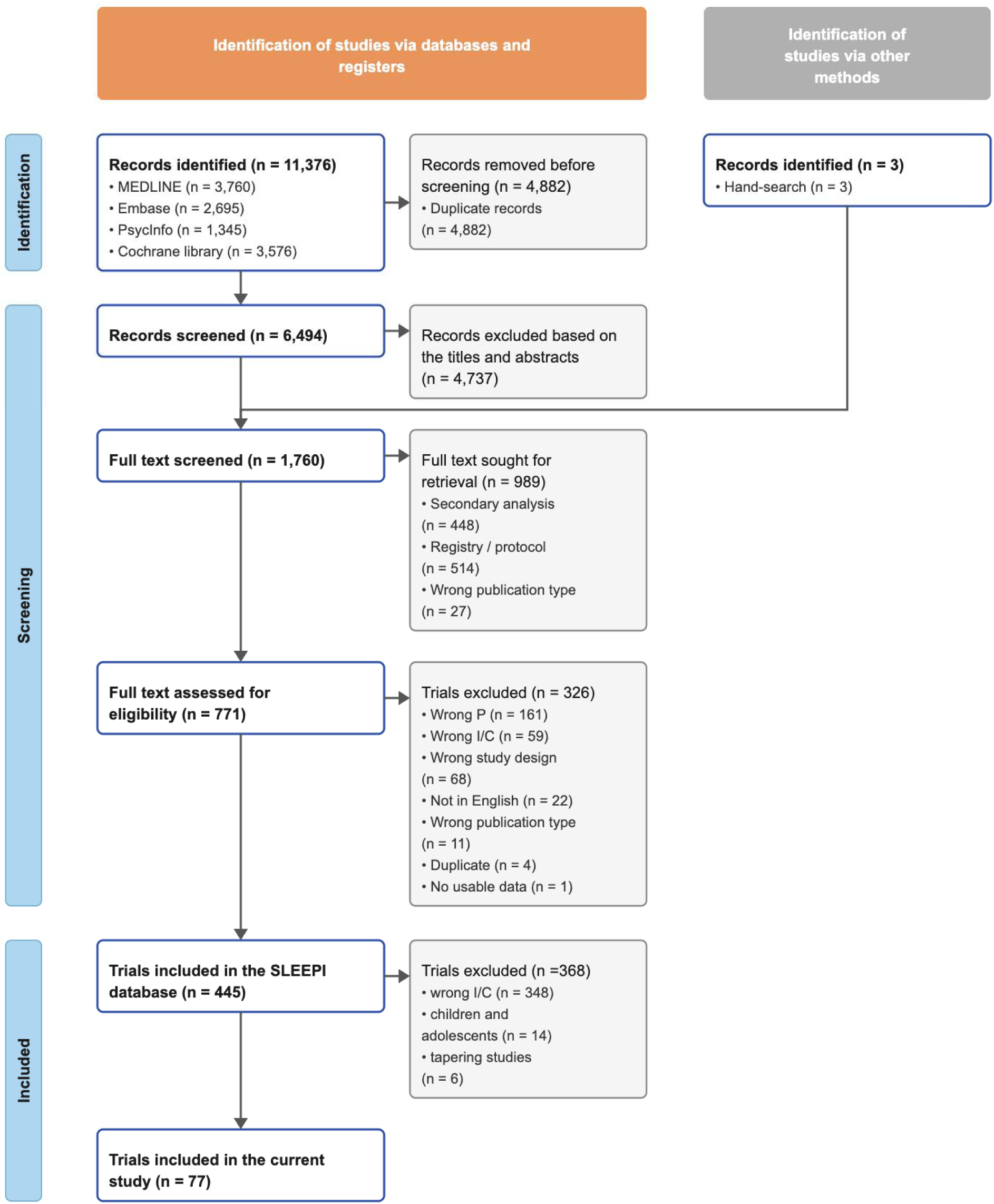
The screening process. I/C = intervention/control; P = population.

**Figure 2.**
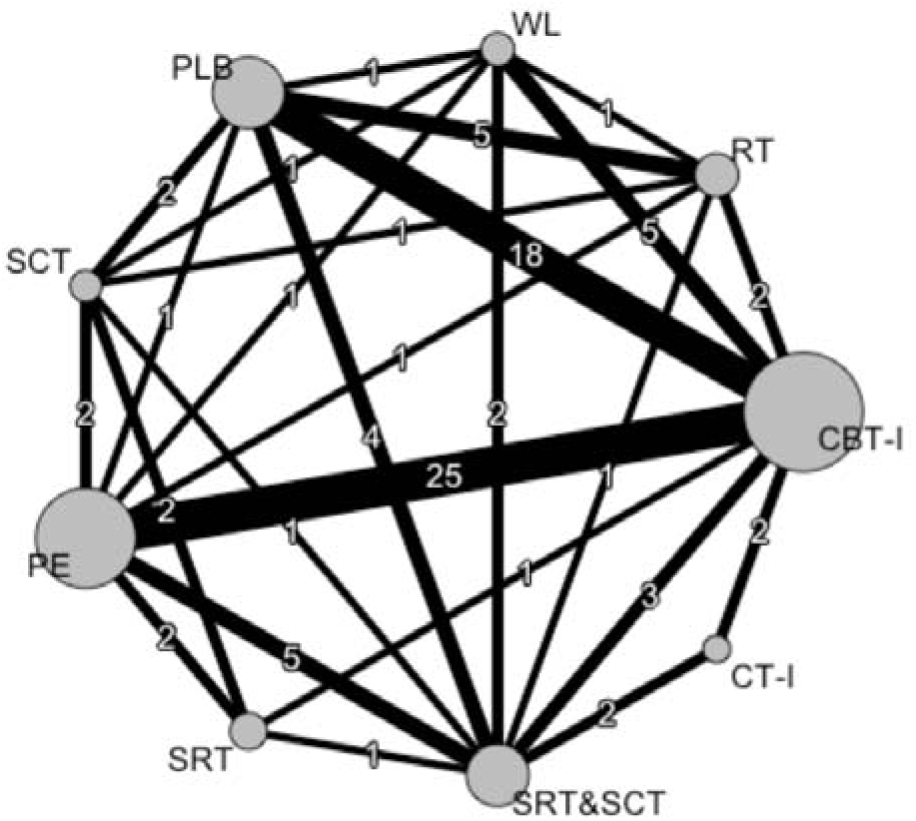
The network diagram. CBT-I = cognitive behavioural therapy for insomnia; CT-I = cognitive therapy for insomnia; PE = psychoeducation (sleep hygiene); PLB = psychological placebo; RT = relaxation therapy; SCT = stimulus control therapy; SRT = sleep restriction therapy; SRT&SCT = sleep restriction therapy and stimulus control therapy; WL = waiting list. The width of lines connecting treatments corresponds to the number of trials performing the corresponding comparison. This number is also given on each line. The size of nodes corresponds to the number of participants randomised to the intervention.

Figure 3 shows the result of the NMA and eAppendix5 shows the results of the pairwise meta-analyses, the direct and indirect estimates of NMA, and the league table. We found that CBT-I, SRT&SCT, SCT and SRT all performed better than all the control conditions (psychoeducation [sleep hygiene], psychological placebo, waitlist, no treatment) and relaxation therapy. There was no evidence of difference among these efficacious interventions. When compared against psychoeducation (sleep hygiene), evidence certainty was moderate for CBT-I, low for SRT&SCT, and very low for SCT and SRT. There was no evidence of difference between psychoeducation and CT-I or relaxation therapy, but CT-I was better than relaxation therapy.

**Figure 3.**
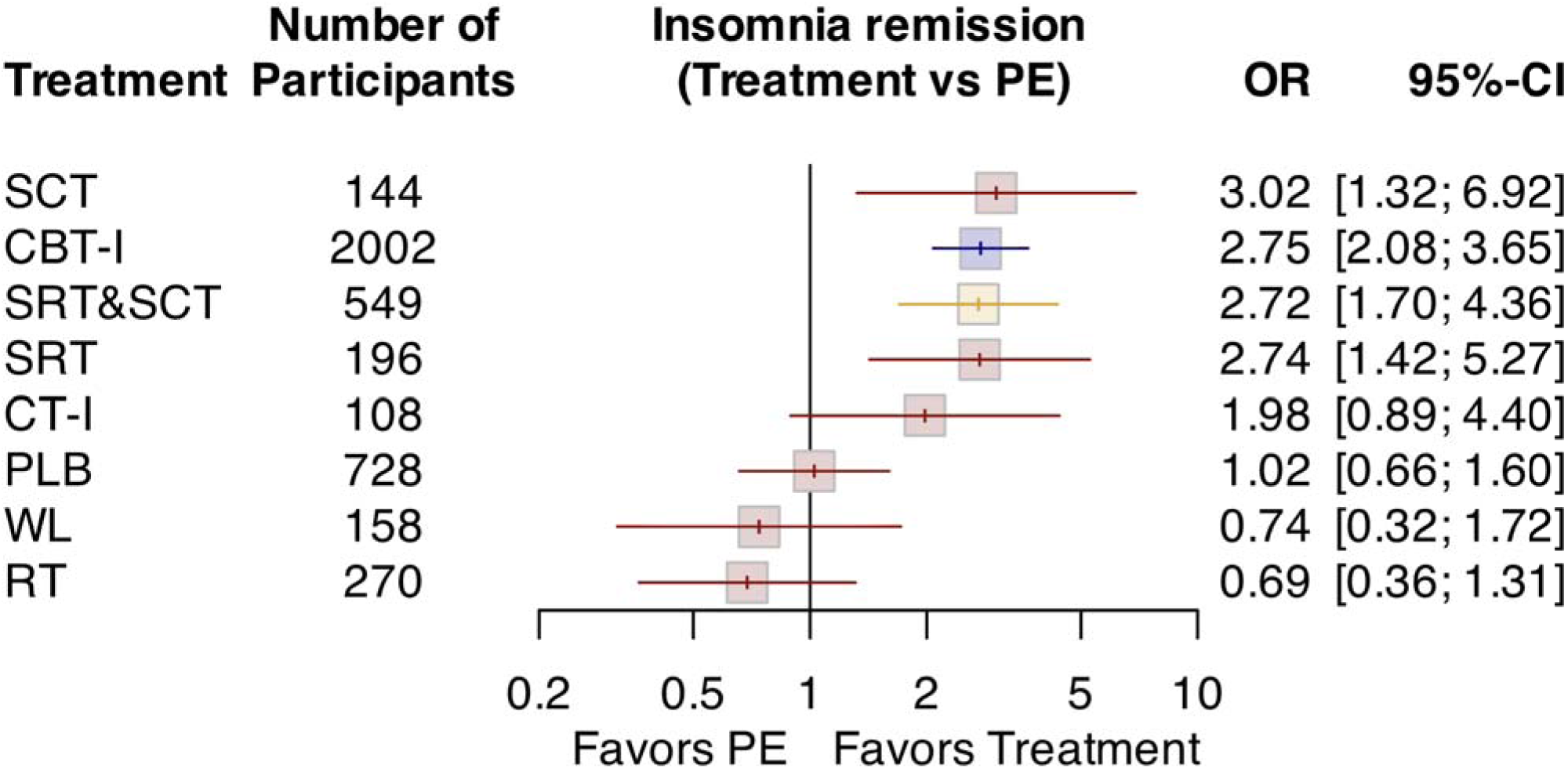
The result of network meta-analysis of insomnia remission, using psychoeducation (sleep hygiene) as the reference. CBT-I = cognitive behavioral therapy for insomnia; CI = confidence interval; CT-I = cognitive therapy for insomnia; NT = no treatment; OR = odds ratio; PE = psychoeducation (sleep hygiene); PLB = psychological placebo; RT = relaxation therapy; SCT = stimulus control therapy; SRT = sleep restriction therapy; SRT&SCT = sleep restriction therapy and stimulus control therapy; WL = waiting list.

Statistical evaluations of transitivity and consistency are presented in eAppendix3. The global design-by-treatment interaction test indicated some inconsistency (p = 0.05). Local assessment by the back-calculation method identified disagreement between direct and indirect evidence in 2 of 26 comparisons, a proportion consistent with empirical expectations.[40]. (eAppendix3) Between-study heterogeneity was moderate (I² = 33.1% [11.2–49.6%]), and the estimated τ² (0.20) was within the range typically observed in published networks.[41] Overall, we interpreted the findings as the assumptions required for network meta-analysis being reasonably met, although sensitivity analyses and cautious interpretations of results were needed.

Figure 4 summarises the primary and secondary outcomes. There was no evidence of difference in dropout. Sleep continuity measures, such as sleep efficiency, sleep latency and wake after sleep onset, were better among SCT, CBT-I, SRT, SRT&SCT, and relaxation therapy. There was no evidence of superiority of cognitive therapy for insomnia or relaxation therapy over psychoeducation in terms of insomnia remission. Weak evidence suggested inferiority of no treatment and waitlist to psychoeducation.

**Figure 4.**
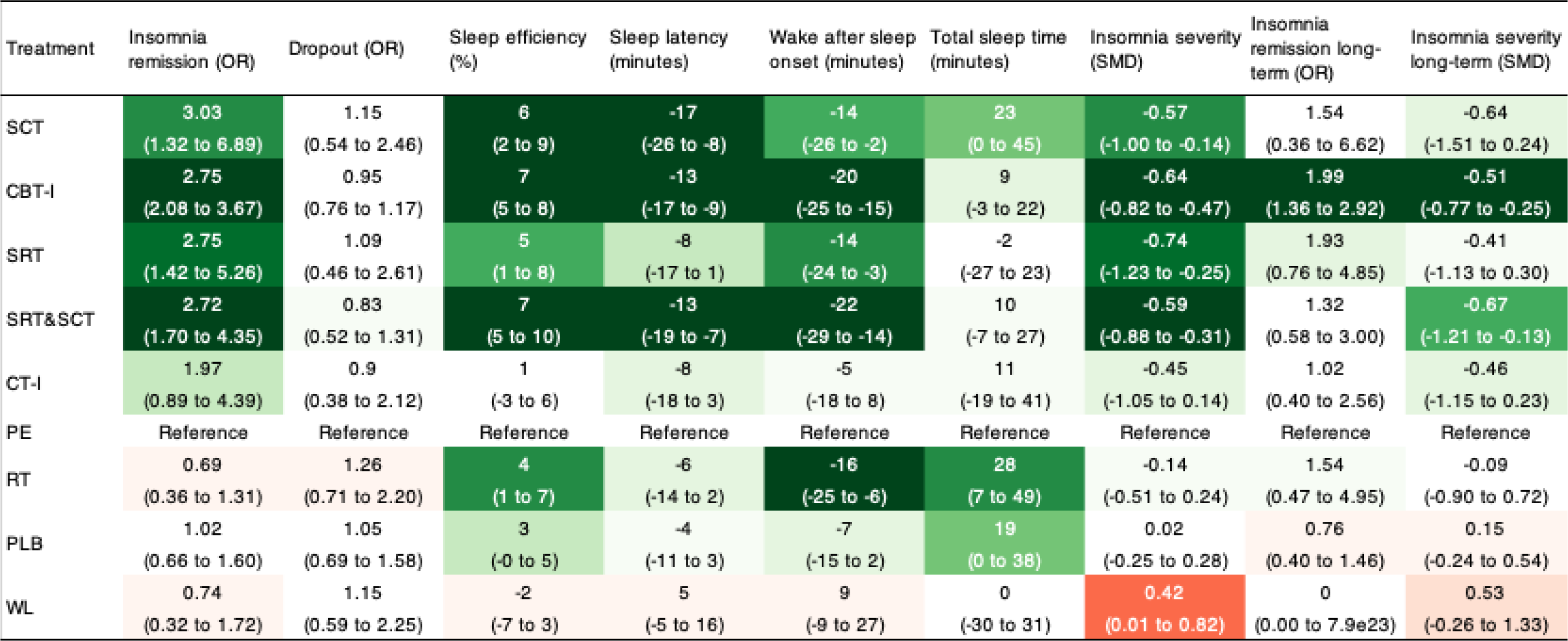
The primary and secondary outcomes. CBT-I = cognitive behavioral therapy for insomnia; CT-I = cognitive therapy for insomnia; NT = no treatment; OR = odds ratio; PE = psychoeducation (sleep hygiene education); PLB = psychological placebo; RT = relaxation therapy; SCT = stimulus control therapy; SMD = standardized mean difference; SRT = sleep restriction therapy; SRT&SCT = sleep restriction therapy and stimulus control therapy; WL = waiting list.

Various a priori and post hoc sensitivity analyses were in line with the primary analysis; the networks were well-connected, and the analyses showed superiority of CBT-I, SRT&SCT, SRT and SCT over psychoeducation.(eAppendix8)

Small-study effects were suspected by the contour-enhanced funnel plots. (eAppendix4) eAppendix6 presents the evaluation of CINeMA. We used the average RoB for the within-study bias domain. We upgraded the within-study bias domain for CBT-I:PE, CBT-I:PLB and CBTI:RT, based on sensitivity analyses excluding high RoB trials. For the reporting bias domain, we set “some concerns” as default, because the lack of preregistration is still prevalent in the field, and visual inspection of funnel plots suggested some reporting bias. We upgraded for those comparing active interventions (CBT-I, SRT&SCT, SRT, SCT, cognitive therapy for insomnia and relaxation therapy). CBT-I was superior to cognitive therapy for insomnia, relaxation therapy and other control conditions with moderate to low confidence in the evidence, SRT&SCT with low to very low confidence, and SRT and SCT mainly with very low confidence.

Given the weighted mean proportion of insomnia remission among sleep hygiene arms of 20%, CBT-I probably leads to remission rate of 41% (95% CI, 34%; 48%), SRT&SCT may lead to remission rate of 40% (30%; 52%), SCT 43% (25%; 63%), and SRT 41% (26%; 57%).

## DISCUSSION

This network meta-analysis compared multicomponent CBT-I with abbreviated behavioural versions while accounting for potential interactions among treatment components. We found that CBT-I increases insomnia remission with moderate confidence in the evidence, whereas SRT&SCT showed similar efficacy with low confidence in the evidence, and SCT alone and SRT alone with very low confidence in the evidence. We found no clear evidence of superiority of CT-I or relaxation therapy over psychoeducation (sleep hygiene education). Relaxation therapy was inferior to CBT-I, SRT&SCT, SCT, SRT, and CT-I.

### Comparisons with other studies and guidelines

Beyond confirming the efficacy of CBT-I, this study adds comparative evidence across treatment packages. Previous pairwise meta-analyses have evaluated SRT, SCT, and other behvioural therapies individually. Current guidelines largely rely on such separate bodies of evidence when making recommendations. By directly comparing multicomponent CBT-I with abbreviated behavioural formats using remission as a common endpoint, our findings provide evidence that simplified behavioural interventions may achieve remission rates comparable to full CBT-I, albeit with lower certainty. This distinction is relevant to treatment selection and service planning.

In general, the results support the American Academy of Sleep Medicine clinical practice guideline.[10,11] We found CBT-I is indeed the most established behavioral and psychological intervention for insomnia. Additionally, our results suggested that SCT+SRT, SCT alone, and SRT alone achieved comparable remission, although the evidence base is limited relative to the full CBT-I. The guideline recommended relaxation therapy, whereas our results did not support its effectiveness. This difference arises from the fact that the guideline interpreted multiple outcomes, while we prioritized the primary outcome (insomnia remission), and from differences in how the very low confidence in the evidence was interpreted.

These findings suggest that while multicomponent CBT-I remains the most established option, simplified behavioural formats may represent pragmatic alternatives when access to comprehensive CBT-I is limited. Previous component-based analyses have identified behavioural elements as central to CBT-I; the present findings extend this evidence by demonstrating that behavioural treatment packages themselves may achieve comparable remission, albeit with lower certainty. Decisions to prioritise abbreviated interventions should therefore balance scalability with the robustness of the supporting evidence.

### Limitations

Our study has several limitations. First, we included only peer-reviewed reports, which may have resulted in the omission of eligible unpublished trials and introduced bias due to missing evidence. The contour-enhanced funnel plots also suggested possible small-study effects or publication bias. However, this approach ensures sufficient detail regarding intervention content and methodological quality.[22] Second, statistical heterogeneity remained despite efforts to enhance comparability by focusing on in-person delivery format. Although sensitivity analyses yielded broadly consistent findings, residual heterogeneity may influence the precision of effect estimates. Third, the certainty of evidence for several abbreviated interventions was low or very low, limiting confidence in their comparative effectiveness. We considered these uncertainties when interpreting the apparent similarity in remission rates between abbreviated formats and full CBT-I. We also included only reports published in English, which may have introduced language bias and limited the representation of evidence from different cultural and healthcare settings.

### Implications for clinical practice and future studies

This study indicated that full-package CBT-I currently has the most robust evidence base to date in terms of both the quantity and quality of available trials and therefore remains the most well-supported intervention for insomnia. However, we found no clear evidence that its effect on remission was superior to those of abbreviated behavioural therapies, particularly sleep restriction therapy and stimulus control therapy. While these behavioural interventions showed similar point estimates for remission, the certainty of evidence was lower than that for full CBT-I. may therefore represent pragmatic alternatives when access to full CBT-I is limited, although their lower certainty of evidence should be considered in treatment and implementation decisions.

In contrast, the clinical benefits of cognitive therapy warrant further evaluation, and relaxation therapy or psychoeducation alone appear insufficient as a stand-alone treatment for insomnia. These findings suggest that behavioral components constitute the central role of CBT-I, while the added value of other components may depend on how they are combined and delivered.

Future research should examine optimal treatment combinations and sequences. In particular, investigating additive benefits of cognitive restructuring with behavioral interventions have the potential to maximise treatment effects. In addition, step-care trial designs—such as initiating treatment with behavioral therapy and subsequently randomizing non-responders to additional or alternative components, including pharmacotherapies—may help clarify the most efficient and clinically relevant treatment pathways.

### Conclusion

Multicomponent CBT-I remains the most robustly supported treatment for insomnia. Abbreviated behavioural formats, particularly sleep restriction therapy and stimulus control therapy, may achieve similar remission rates, although with lower certainty of evidence. In settings where access to full CBT-I is constrained, simplified behavioural interventions may represent pragmatic alternatives. Further research should determine how best to sequence and integrate these approaches within routine care.

## Data Availability

All data analysed in this study were extracted from published reports. The extracted aggregate dataset and analysis code supporting the findings are available from the corresponding author upon reasonable request.

## Ethics statements

Ethical approval was not required.

## Acknowledgements

The views expressed are those of the authors and not necessarily those of affiliated organizations. We thank Ralph de Vries for his contribution to the systematic literature search.

## Registration

This protocol is prospectively registered in the Open Science Framework (https://osf.io/z48r2/).

## Contributions of authors

Conceptualization: MS, SK, MI, RT, HNT, SH, RY, SN, HN, KI, SS, DN, YT, KK, SSh, AI, MP, AVS, YF; Data curation: MS, SK, MI, RT, HNT, SH, RY, SN, HN, KI, SS, DN, YT, KK, SSh, AI, YF; Formal Analysis: YF; Funding acquisition: YF; Investigation: MS, SK, MI, RT, HNT, SH, RY, SN, HN, KI, SS, DN, YT, KK, SSh, AI, MP, AVS, YF; Methodology: MS, YF; Project administration: YF; Resources:; Software: YF; Supervision: MP, AVS; Validation: MS; Visualization: YF; Writing – original draft: MS, YF; Writing – review & editing: MS, SK, MI, RT, HNT, SH, RY, SN, HN, KI, SS, DN, YT, KK, SSh, AI, MP, AVS, YF

## Support

This study was supported partly by a grant from SENSHIN Medical Research Foundation given to YF

## Declarations of interest

MS is employed YF has authored multiple academic papers, books, and articles on cognitive behavioral therapy for insomnia and has received honoraria for lectures and supervisory roles.

SK, CT, EH, SH, HN and AI declare no conflicts of interest.

HNT is employed as Director and Principal Researcher at Awarefy Inc., a company that develops and commercializes a CBT-based digital mental health application.

RY has authored multiple academic papers, books, and articles on cognitive behavioral therapy for insomnia and the Sleep Research Institute of Edogawa University, to which Ry belongs, is collaborating with PARAMOUNT BED CO., LTD. and receiving research funding.

SN has received research funding and donations from Amazon, Tsukuba City, Nomura Real Estate, S’UIMIN Inc., and Mitsubishi Electric.

## Notes

### Competing Interest Statement

MS is employed in the Department of Neurodevelopmental Medicine, Nagoya City University Graduate School of Medical Sciences, which is an endowment department supported by the City of Nagoya, and has received a personal fee from Daiichi-Sankyo outside the submitted work. MS and YF has authored multiple academic papers, books, and articles on cognitive behavioral therapy for insomnia and has received honoraria for lectures and supervisory roles.
SK, CT, EH, SH, HN and AI declare no conflicts of interest.
HNT is employed as Director and Principal Researcher at Awarefy Inc., a company that develops and commercializes a CBT-based digital mental health application.
RY has authored multiple academic papers, books, and articles on cognitive behavioral therapy for insomnia and the Sleep Research Institute of Edogawa University, to which Ry belongs, is collaborating with PARAMOUNT BED CO., LTD. and receiving research funding.
SN has received research funding and donations from Amazon, Tsukuba City, Nomura Real Estate, S'UIMIN Inc., and Mitsubishi Electric.

### Clinical Protocols

https://osf.io/z48r2/

### Author Declarations

This systematic review and network meta-analysis used only aggregate human data reported in previously published peer-reviewed articles and their supplementary materials. All source data were publicly available before the initiation of the study. No individual-level, non-public, or author-supplied data were used.

