## SUPPLEMENTARY APPENDIX for "Comparative efficacy and acceptability of cognitive-behavioural therapy for insomnia and its abbreviated versions: a systematic review and network meta-analysis"

This preprint supplement provides the full search strategies, aggregate diagnostic analyses, network estimates, certainty assessments, secondary outcomes, sensitivity analyses, and the PRISMA-NMA checklist. Detailed study-level extraction tables and study-level risk-of-bias assessments are available from the authors upon reasonable request.

### 1. SEARCH STRATEGIES

**Ovid Medline Search Results – 15 Dec 2025**

| **Search** | **Query** | **Results** |
| --- | --- | --- |
| #5 | **3 and 4** | 3,760 |
| #4 | **(("Clinical Trial" or "Controlled Clinical Trial").pt. or "Controlled Clinical Trials as Topic"/ or exp "Randomized Controlled Trials as Topic"/ or exp "Randomized Controlled Trial"/ or "Randomized Controlled Trial".pt. or ("random*" or "RCT*").ti,kf. or "randomly".ab. or (("pragmatic" or "practical") adj "clinical trial*").ti,ab,kf. or (("non-inferiority" or "noninferiority" or "superiority" or "equivalence") adj3 "trial*").ti,ab,kf. or ("control*" adj3 ("group*" or "wait*")).ti,ab,kf.) not ("comment"/ or "editorial"/ or "letter"/ or ((exp "animals"/ or exp "models, animal"/) not "humans"/))** | 1,876,486 |
| #3 | **1 and 2** | 16,174 |
| #2 | **"Psychotherapy"/ or exp "Behavior Therapy"/ or exp "Sleep Hygiene"/ or exp "Relaxation Therapy"/ or "Bibliotherapy"/ or (("behavio*" adj3 ("therap*" or "intervent*")) or ("cognit*" adj3 ("therap*" or "intervent*")) or ("behavio*" adj3 "psychotherap*") or ("cognit*" adj3 "psychotherap*") or "bibliotherap*" or "cognitive restructur*" or "cognitive refram*" or "cognitive reapprais*" or "third wave" or "3rd wave" or "mindfulness" or ("sleep*" adj3 ("restrict*" or "depriv*" or "consol*")) or "time-in-bed restrict*" or "bedtime restrict*" or "Acceptance and Commitment" or ("stimulus" adj3 "control*") or "sleep hygiene" or "sleep education" or "sleep diar*" or "constructive worr*" or "imagery rehearsal" or "relaxation" or "paradoxical intent*" or "CBT" or "CBTI" or "CBT-I").ab,ti,kf.** | 395,821 |
| #1 | **"Sleep Wake Disorders"/ or exp "Sleep Initiation and Maintenance Disorders"/ or "Wakefulness"/ or ("insomni*" or "sleepless*" or "hyposomn*" or "hypo-somn*" or "early awake*" or ("sleep initiation" adj3 ("dysfunction*" or "disorder*")) or ("sleep*" adj3 "disturb*") or "wakeful*" or "subwakeful*" or "wake state*" or "waking state*" or "early morning awakening" or "nocturnal awakening" or "sleep state*" or "sleeping state*").ab,ti,kf.** | 118,098 |

**Embase.com Search Results – 15 Dec 2025**

| **Search** | **Query** | **Results** |
| --- | --- | --- |
| #6 | **#5 NOT ('conference abstract'/it OR 'conference review'/it) NOT 'clinical trial':dtype** | 2,695 |
| #5 | **#3 AND #4** | 5,450 |
| #4 | **('clinical trial (topic)'/exp OR 'randomized controlled trial'/exp OR ("random*" OR "RCT*"):ti,kw OR ("randomly"):ab OR (("pragmatic" OR "practical") NEAR "clinical trial*"):ab,ti,kw OR (("non-inferiority" OR "noninferiority" OR "superiority" OR "equivalence") NEAR/3 "trial*"):ab,ti,kw OR ("control*" NEAR/3 ("group*" or "wait*")):ab,ti,kw) NOT ("editorial"/exp OR "letter"/exp OR (("animal"/exp OR "animal model"/exp OR 'nonhuman'/exp) NOT "human"/exp))** | 2,544,614 |
| #3 | **#1 AND #2** | 19,695 |
| #2 | **'psychotherapy'/de OR 'behavior therapy'/exp OR 'cognitive therapy'/exp OR 'cognitive behavioral therapy for insomnia'/exp OR 'sleep hygiene'/exp OR 'sleep education'/exp OR 'sleep diary'/exp OR 'imagery rehearsal therapy'/exp OR 'relaxation training'/exp OR 'paradoxical intention'/exp OR 'bibliotherapy'/exp OR 'sleep deprivation'/exp OR 'sleep restriction therapy'/exp OR 'cognitive reappraisal'/exp OR 'mindfulness'/exp OR 'stimulus control'/exp OR 'stimulus control therapy'/exp OR (("behavio*" NEAR/3 ("therap*" OR "intervent*")) OR ("cognit*" NEAR/3 ("therap*" OR "intervent*")) OR ("behavio*" NEAR/3 "psychotherap*") OR ("cognit*" NEAR/3 "psychotherap*") OR "bibliotherap*" OR "cognitive restructur*" OR "cognitive refram*" OR "cognitive reapprais*" OR "third wave" OR "3rd wave" OR "mindfulness" OR ("sleep*" NEAR/3 ("restrict*" OR "depriv*" OR "consol*")) OR "time-in-bed restrict*" OR "bedtime restrict*" OR "Acceptance and Commitment" OR ("stimulus" NEAR/3 "control*") OR "sleep hygiene" OR "sleep education" OR "sleep diar*" OR "constructive worr*" OR "imagery rehearsal" OR "relaxation" OR "paradoxical intent*" OR "CBT" OR "CBTI" OR "CBT-I"):ab,ti,kw** | 548,531 |
| #1 | **'sleep disorder'/de OR 'insomnia'/exp OR 'wakefulness'/exp OR 'wakefulness after sleep onset'/exp ("insomni*" OR "sleepless*" OR "hyposomn*" OR "hypo-somn*" OR "early awake*" OR ("sleep initiation" NEAR/3 ("dysfunction*" OR "disorder*")) OR ("sleep*" NEAR/3 "disturb*") OR wakeful* OR "subwakeful*" OR "wake state*" OR "waking state*" OR "early morning awakening" OR "nocturnal awakening" OR "sleep state*" OR "sleeping state*"):ab,ti,kw** | 105,036 |

**APA PsycInfo (Ebsco) Search Results – 15 Dec 2025**

| **Search** | **Query** | **Results** |
| --- | --- | --- |
| S5 | **S3 AND S4** | 1,345 |
| S4 | **(DE "Clinical Trials" OR DE "Randomized Controlled Trials" OR DE "Randomized Clinical Trials" OR TI ("random*" OR "RCT*") OR AB ("randomly") OR KW ("random*" OR "RCT*") OR TI (("pragmatic" OR "practical") N0 "clinical trial*") OR AB (("pragmatic" OR "practical") N0 "clinical trial*") OR KW (("pragmatic" OR "practical") N0 "clinical trial*") OR TI ((("non-inferiority" OR "noninferiority" OR "superiority" OR "equivalence") N3 "trial*") OR ("control*" N3 ("group*" OR "wait*"))) OR AB ((("non-inferiority" OR "noninferiority" OR "superiority" OR "equivalence") N3 "trial*") OR ("control*" N3 ("group*" OR "wait*"))) OR KW ((("non-inferiority" OR "noninferiority" OR "superiority" OR "equivalence") N3 "trial*") OR ("control*" N3 ("group*" OR "wait*")))) NOT (PO "Animal" NOT PO "Human")** | 235,892 |
| S3 | **S1 AND S2** | 9,769 |
| S2 | **DE "Psychotherapy" OR DE "Individual Psychotherapy" OR DE "Behavior Therapy" OR DE "Cognitive Therapy" OR DE "Cognitive Behavior Therapy" OR DE "Mindfulness-Based Cognitive Therapy" OR DE "Mindfulness" OR DE "Relaxation Therapy" OR DE "Progressive Relaxation Therapy" OR DE "Paradoxical Techniques" OR DE "Bibliotherapy" OR DE "Sleep Deprivation" OR DE "Stimulus Control" OR TI (("behavio*" N3 ("therap*" OR "intervent*")) OR ("cognit*" N3 ("therap*" OR "intervent*")) OR ("behavio*" N3 "psychotherap*") OR ("cognit*" N3 "psychotherap*") OR "bibliotherap*" OR "cognitive restructur*" OR "cognitive refram*" OR "cognitive reapprais*" OR "third wave" OR "3rd wave" OR "mindfulness" OR ("sleep*" N3 ("restrict*" OR "depriv*" OR "consol*")) OR "time-in-bed restrict*" OR "bedtime restrict*" OR "Acceptance and Commitment" OR ("stimulus" N3 "control*") OR "sleep hygiene" OR "sleep education" OR "sleep diar*" OR "constructive worr*" OR "imagery rehearsal" OR "relaxation" OR "paradoxical intent*" OR "CBT" OR "CBTI" OR "CBT-I") OR AB (("behavio*" N3 ("therap*" OR "intervent*")) OR ("cognit*" N3 ("therap*" OR "intervent*")) OR ("behavio*" N3 "psychotherap*") OR ("cognit*" N3 "psychotherap*") OR "bibliotherap*" OR "cognitive restructur*" OR "cognitive refram*" OR "cognitive reapprais*" OR "third wave" OR "3rd wave" OR "mindfulness" OR ("sleep*" N3 ("restrict*" OR "depriv*" OR "consol*")) OR "time-in-bed restrict*" OR "bedtime restrict*" OR "Acceptance and Commitment" OR ("stimulus" N3 "control*") OR "sleep hygiene" OR "sleep education" OR "sleep diar*" OR "constructive worr*" OR "imagery rehearsal" OR "relaxation" OR "paradoxical intent*" OR "CBT" OR "CBTI" OR "CBT-I") OR KW (("behavio*" N3 ("therap*" OR "intervent*")) OR ("cognit*" N3 ("therap*" OR "intervent*")) OR ("behavio*" N3 "psychotherap*") OR ("cognit*" N3 "psychotherap*") OR "bibliotherap*" OR "cognitive restructur*" OR "cognitive refram*" OR "cognitive reapprais*" OR "third wave" OR "3rd wave" OR "mindfulness" OR ("sleep*" N3 ("restrict*" OR "depriv*" OR "consol*")) OR "time-in-bed restrict*" OR "bedtime restrict*" OR "Acceptance and Commitment" OR ("stimulus" N3 "control*") OR "sleep hygiene" OR "sleep education" OR "sleep diar*" OR "constructive worr*" OR "imagery rehearsal" OR "relaxation" OR "paradoxical intent*" OR "CBT" OR "CBTI" OR "CBT-I")** | 267,567 |
| S1 | **DE "Sleep Wake Disorders" OR DE "Insomnia" OR DE "Wakefulness" OR TI ("insomni*" OR "sleepless*" OR "hyposomn*" OR "hypo-somn*" OR "early awake*" OR ("sleep initiation" N3 ("dysfunction*" OR "disorder*")) OR ("sleep*" N3 "disturb*") OR "wakeful*" OR "subwakeful*" OR "wake state*" OR "waking state*" OR "early morning awakening" OR "nocturnal awakening" OR "sleep state*" OR "sleeping state*") OR AB ("insomni*" OR "sleepless*" OR "hyposomn*" OR "hypo-somn*" OR "early awake*" OR ("sleep initiation" N3 ("dysfunction*" OR "disorder*")) OR ("sleep*" N3 "disturb*") OR "wakeful*" OR "subwakeful*" OR "wake state*" OR "waking state*" OR "early morning awakening" OR "nocturnal awakening" OR "sleep state*" OR "sleeping state*") OR KW ("insomni*" OR "sleepless*" OR "hyposomn*" OR "hypo-somn*" OR "early awake*" OR ("sleep initiation" N3 ("dysfunction*" OR "disorder*")) OR ("sleep*" N3 "disturb*") OR "wakeful*" OR "subwakeful*" OR "wake state*" OR "waking state*" OR "early morning awakening" OR "nocturnal awakening" OR "sleep state*" OR "sleeping state*")** | 52,544 |

**Cochrane Library (Wiley) Search Results – 15 Dec 2025**

| **Search** | **Query** | **Results** |
| --- | --- | --- |
| #6 | **#5 NOT conference:pt** | 3,576 |
| #5 | **#3 AND #4** | 4,740 |
| #4 | **(random* OR RCT*):ti,kw OR (randomly):ab OR ((pragmatic OR practical) NEXT (clinical NEXT trial*)):ab,ti,kw OR ((non-inferiority OR noninferiority OR superiority OR equivalence) NEAR/3 trial*):ab,ti,kw OR (control* NEAR/3 (group* OR wait*)):ab,ti,kw** | 1,253,846 |
| #3 | **#1 AND #2** | 6,792 |
| #2 | **((behavio* NEAR/3 (therap* OR intervent*)) OR (cognit* NEAR/3 (therap* OR intervent*)) OR (behavio* NEAR/3 psychotherap*) OR (cognit* NEAR/3 psychotherap*) OR bibliotherap* OR (cognitive NEXT restructur*) OR (cognitive NEXT refram*) OR (cognitive NEXT reapprais*) OR (third NEXT wave) OR (3rd NEXT wave) OR mindfulness OR (sleep* NEAR/3 (restrict* OR depriv* OR consol*)) OR (time NEXT in NEXT bed NEXT restrict*) OR (bedtime NEXT restrict*) OR (Acceptance NEAR/3 Commitment) OR (stimulus NEAR/3 control*) OR (sleep NEXT hygiene) OR (sleep NEXT education) OR (sleep NEXT diar*) OR (constructive NEXT worr*) OR (imagery NEXT rehearsal) OR relaxation OR (paradoxical NEXT intent*) OR CBT OR CBTI OR CBT-I):ab,ti,kw** | 97,193 |
| #1 | **(insomni* OR sleepless* OR hyposomn* OR (hypo NEXT somn*) OR (early NEXT awake*) OR ((sleep NEXT initiation) NEAR/3 (dysfunction* OR disorder*)) OR (sleep* NEAR/3 disturb*) OR wakeful* OR subwakeful* OR (wake NEXT state*) OR (waking NEXT state*) OR (early NEXT morning NEXT awakening) OR (nocturnal NEXT awakening) OR (sleep NEXT state*) OR (sleeping NEXT state*)):ab,ti,kw** | 27,730 |

### 2. ASSESSMENT OF TRANSITIVITY

#### Box plots

\# Mean age

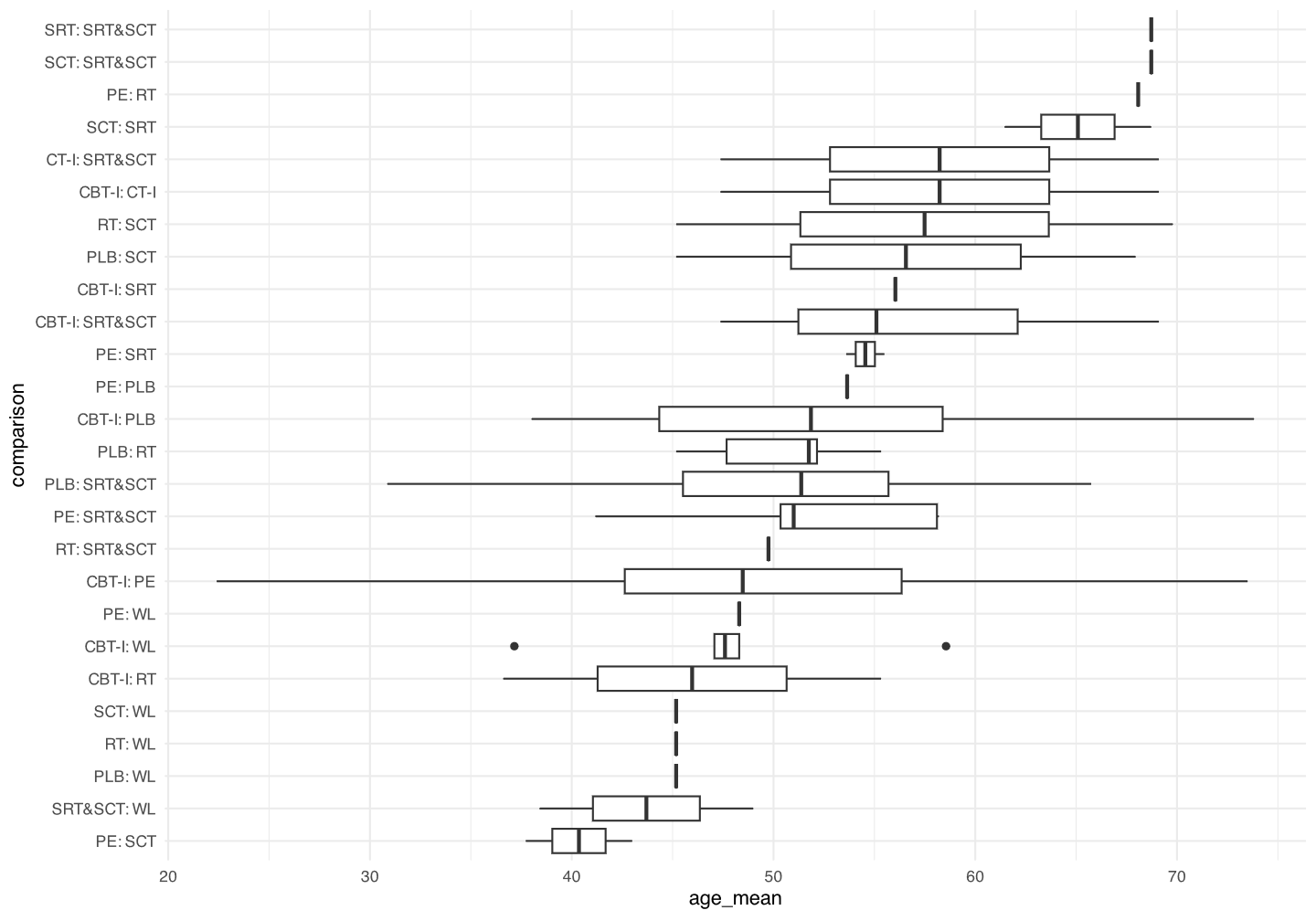

\# Proportion of females

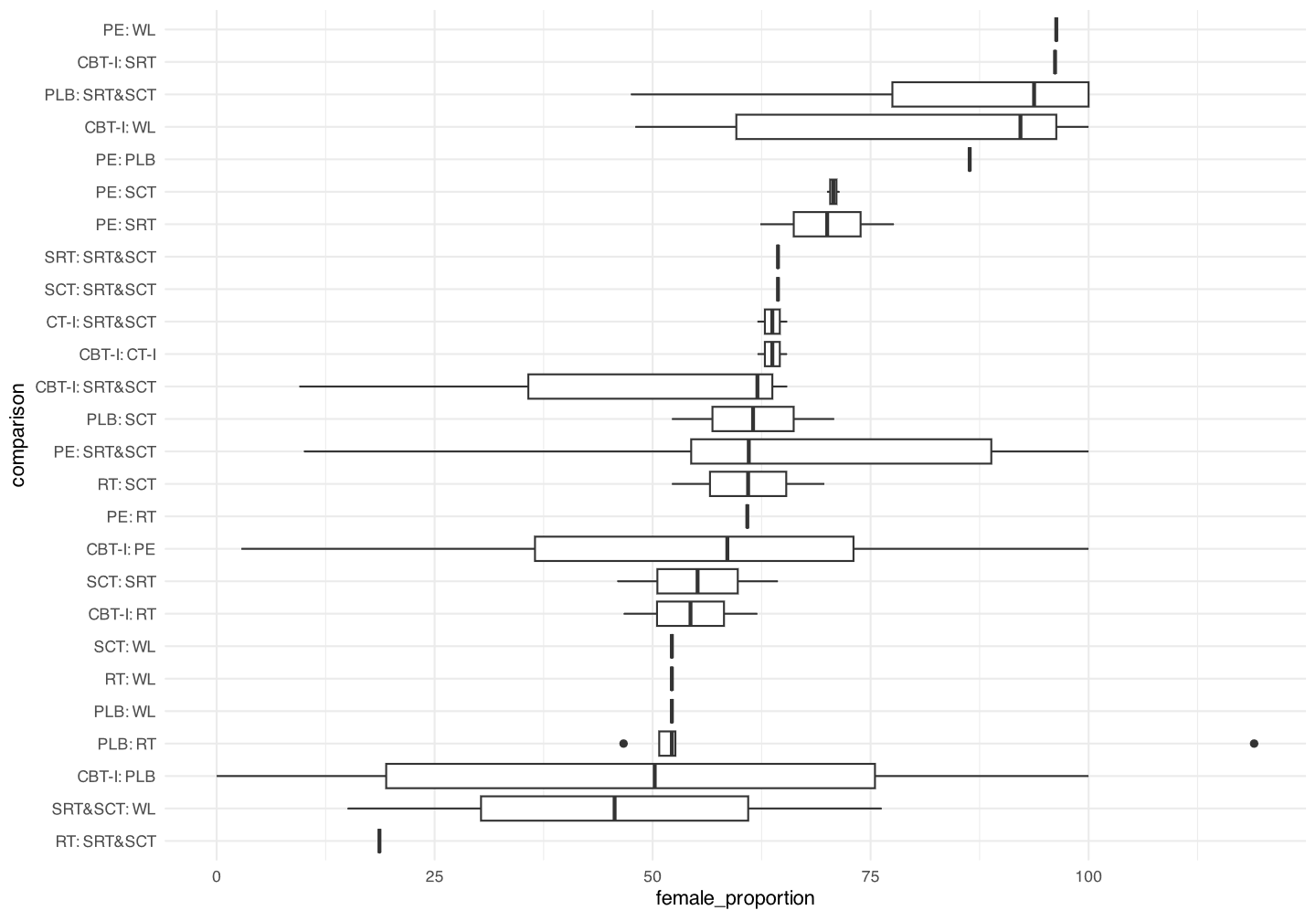

\# Baseline severity

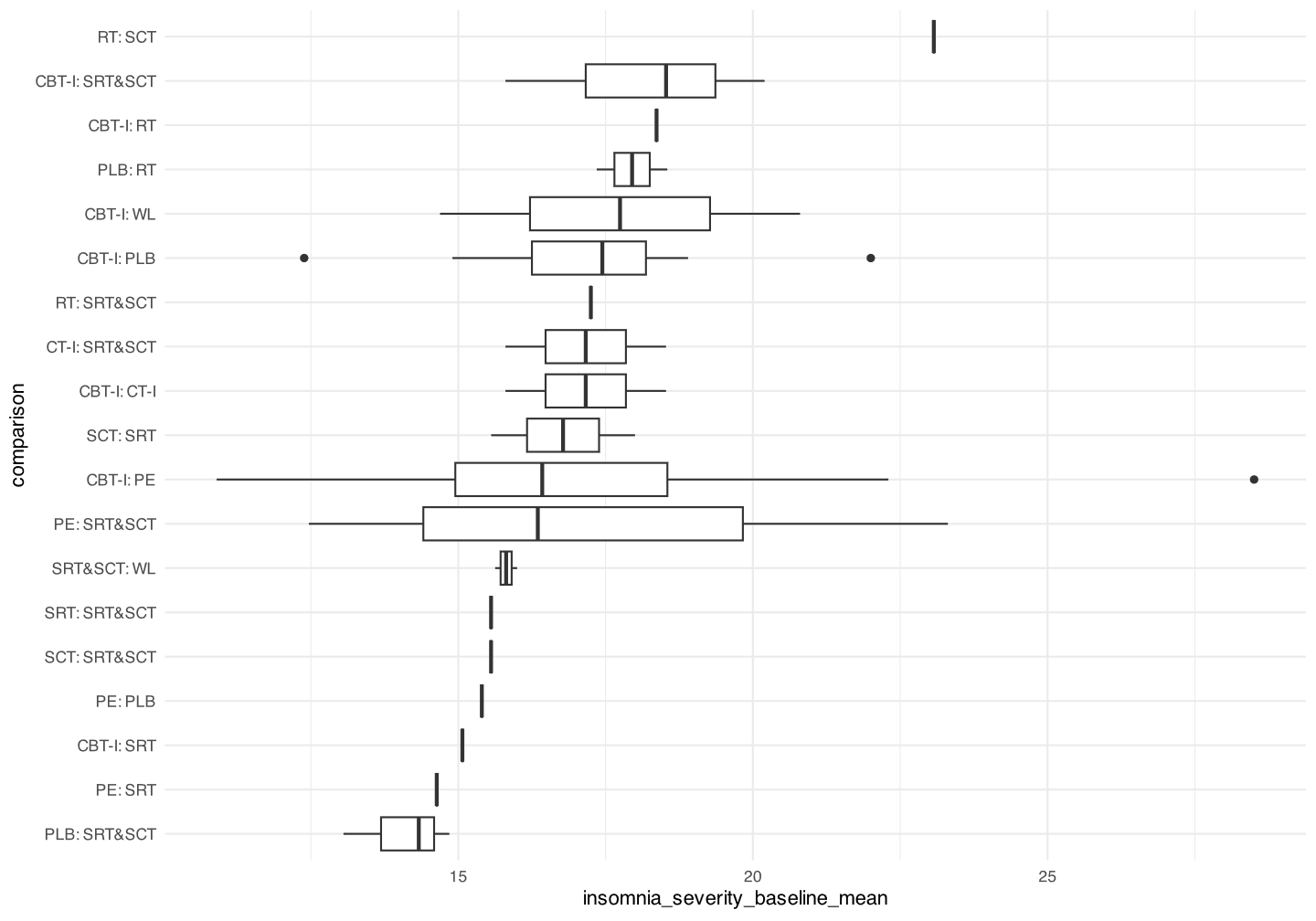

#### Global (design-by-treatment) test

```
Q statistics to assess homogeneity / consistency

                     Q df p-value
Total           113.58 76  0.0034
Within designs   71.97 56  0.0739
Between designs  41.61 20  0.0031

Design-specific decomposition of within-designs Q statistic

             Design     Q df p-value
 CBT-I:CT-I:SRT&SCT 10.63  2  0.0049
         PE:SRT&SCT 12.52  4  0.0139
             PLB:RT  8.52  2  0.0141
        PLB:SRT&SCT  5.53  3  0.1368
           PE:CBT-I 27.42 23  0.2383
             PE:SRT  0.12  1  0.7310
         SRT&SCT:WL  0.06  1  0.8081
             PE:SCT  0.03  1  0.8703
          CBT-I:PLB  6.96 16  0.9740
           CBT-I:WL  0.17  3  0.9820

Between-designs Q statistic after detaching of single designs
(influential designs have p-value markedly different from 0.0031)

 Detached design     Q df p-value
     PE:CBT-I:WL 34.07 18  0.0123
 SCT:SRT:SRT&SCT 34.83 18  0.0099
         SCT:SRT 36.24 19  0.0099
          PE:SRT 37.02 19  0.0079
   PLB:RT:SCT:WL 34.70 17  0.0068
       CBT-I:SRT 38.64 19  0.0049
     PLB:SRT&SCT 38.71 19  0.0048
           PE:RT 38.73 19  0.0048
        CBT-I:RT 38.93 19  0.0045
          PE:PLB 39.13 19  0.0042
        PE:CBT-I 39.37 19  0.0040
    CBT-I:PLB:RT 38.21 18  0.0036
        CBT-I:WL 39.75 19  0.0035
   CBT-I:SRT&SCT 40.31 19  0.0030
          PE:SCT 40.88 19  0.0025
       CBT-I:PLB 41.10 19  0.0023
      SRT&SCT:WL 41.32 19  0.0022
          PLB:RT 41.46 19  0.0021
         PLB:SCT 41.54 19  0.0020
      RT:SRT&SCT 41.54 19  0.0020
      PE:SRT&SCT 41.60 19  0.0020

Q statistic to assess consistency under the assumption of
a full design-by-treatment interaction random effects model

                    Q df p-value tau.within tau2.within
Between designs 31.43 20  0.0498     0.3410      0.1163
```

#### Local (back-calculation) test

```
Separate indirect from direct evidence (SIDE) using back-calculation method

Random effects model:

    comparison  k prop    nma  direct indir.    RoR     z p-value
    CBT-I:CT-I  2 0.83 1.3923  1.4528 1.1392 1.2753  0.24  0.8126
      CBT-I:PE 25 0.84 2.7548  2.5165 4.4816 0.5615 -1.46  0.1451
     CBT-I:PLB 18 0.73 2.6878  3.0589 1.8998 1.6101  1.10  0.2702
      CBT-I:RT  2 0.32 4.0017  6.0242 3.3062 1.8221  0.88  0.3766
     CBT-I:SRT  1 0.31 1.0055  1.8758 0.7619 2.4619  1.24  0.2140
 CBT-I:SRT&SCT  3 0.35 1.0131  0.9339 1.0578 0.8829 -0.26  0.7912
      CBT-I:WL  5 0.62 3.7282  2.7248 6.2605 0.4352 -0.97  0.3317
  CT-I:SRT&SCT  2 0.81 0.7276  0.7531 0.6257 1.2037  0.18  0.8537
        PLB:PE  1 0.04 1.0249  4.5000 0.9597 4.6887  1.37  0.1701
         RT:PE  1 0.23 0.6884  1.4066 0.5591 2.5158  1.18  0.2398
        SCT:PE  2 0.12 3.0226  6.0474 2.7572 2.1933  0.60  0.5499
        SRT:PE  2 0.43 2.7396  4.2459 1.9636 2.1623  1.14  0.2531
    SRT&SCT:PE  5 0.25 2.7192  2.3979 2.8372 0.8452 -0.30  0.7612
         WL:PE  1 0.17 0.7389  0.0855 1.1466 0.0745 -2.26  0.0235
        PLB:RT  5 0.48 1.4889  1.2104 1.7971 0.6735 -0.61  0.5429
       PLB:SCT  2 0.28 0.3391  0.1387 0.4809 0.2885 -1.30  0.1926
   PLB:SRT&SCT  4 0.39 0.3769  0.5288 0.3034 1.7428  1.10  0.2699
        PLB:WL  1 0.11 1.3871  2.0000 1.3274 1.5067  0.29  0.7751
        RT:SCT  1 0.16 0.2277  0.0438 0.3112 0.1406 -1.46  0.1430
    RT:SRT&SCT  1 0.26 0.2532  0.2574 0.2517 1.0228  0.03  0.9768
         RT:WL  1 0.11 0.9317  0.9375 0.9309 1.0070  0.00  0.9965
       SCT:SRT  2 0.62 1.1033  0.7300 2.1572 0.3384 -1.20  0.2312
   SCT:SRT&SCT  1 0.42 1.1116  0.5208 1.9306 0.2698 -1.52  0.1274
        SCT:WL  1 0.20 4.0907 21.4286 2.6734 8.0155  1.51  0.1313
   SRT:SRT&SCT  1 0.29 1.0075  0.3472 1.5572 0.2230 -1.87  0.0620
    SRT&SCT:WL  2 0.29 3.6801  2.4672 4.3183 0.5713 -0.58  0.5639

Legend:
 comparison - Treatment comparison
 k          - Number of studies providing direct evidence
 prop       - Direct evidence proportion
 nma        - Estimated treatment effect (OR) in network meta-analysis
 direct     - Estimated treatment effect (OR) derived from direct evidence
 indir.     - Estimated treatment effect (OR) derived from indirect evidence
 RoR        - Ratio of Ratios (direct versus indirect)
 z          - z-value of test for disagreement (direct versus indirect)
 p-value    - p-value of test for disagreement (direct versus indirect)
```

### 3. ASSESSMENT OF PUBLICATION BIAS AND SMALL STUDY EFFECTS

**4.1. Insomnia remission (dichotomous)**
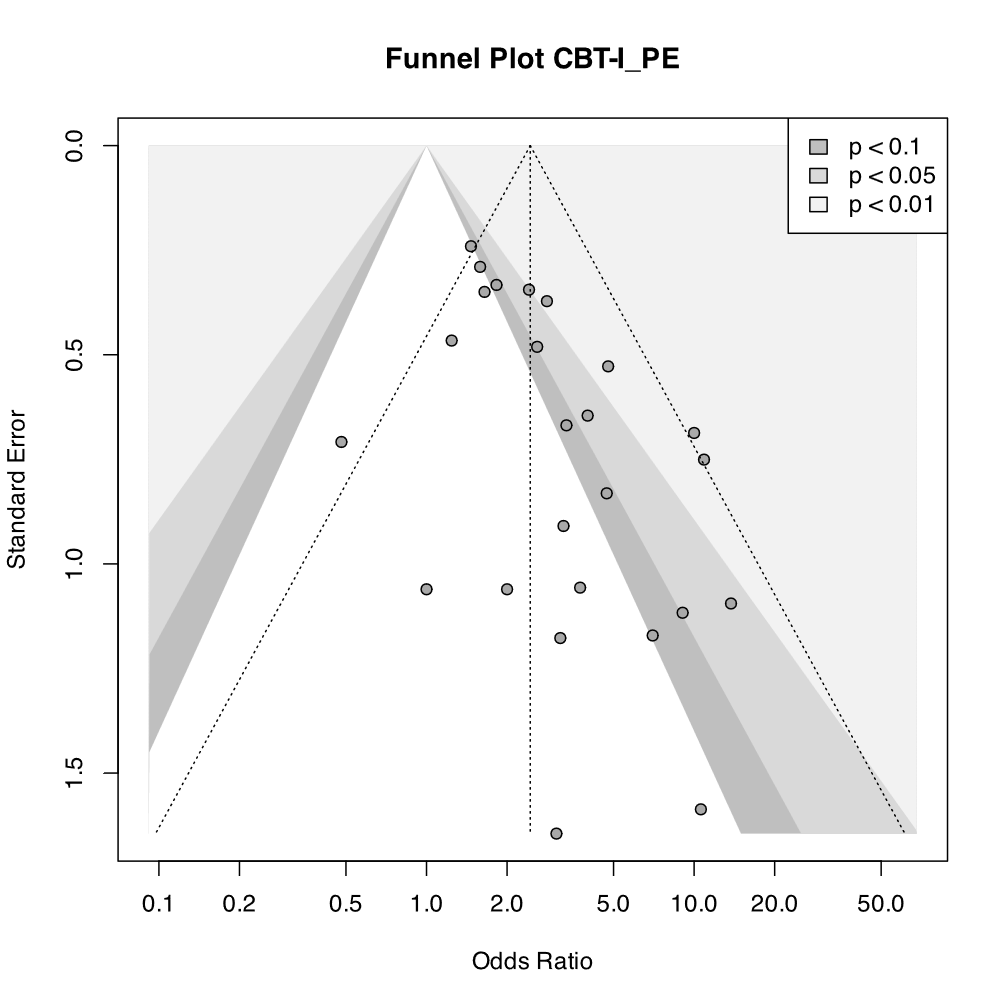

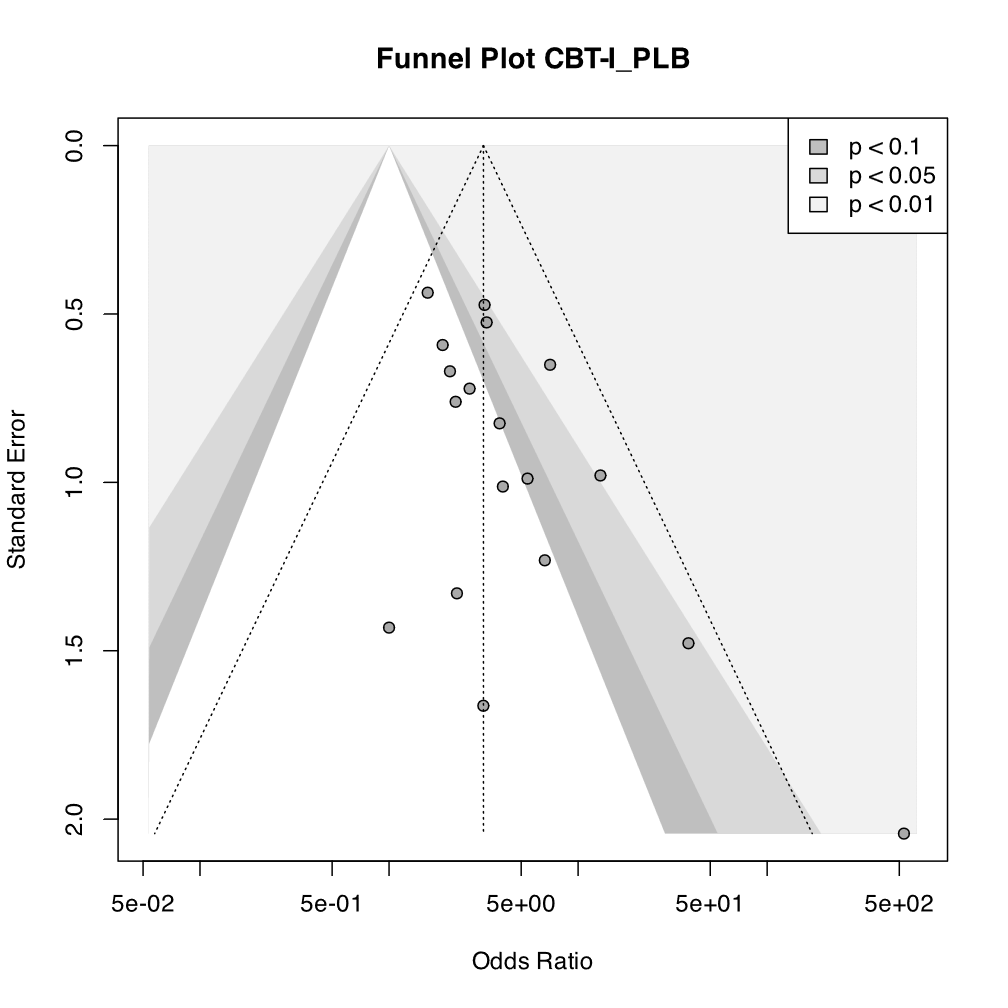
 **4.2. Insomnia severity (continuous)**

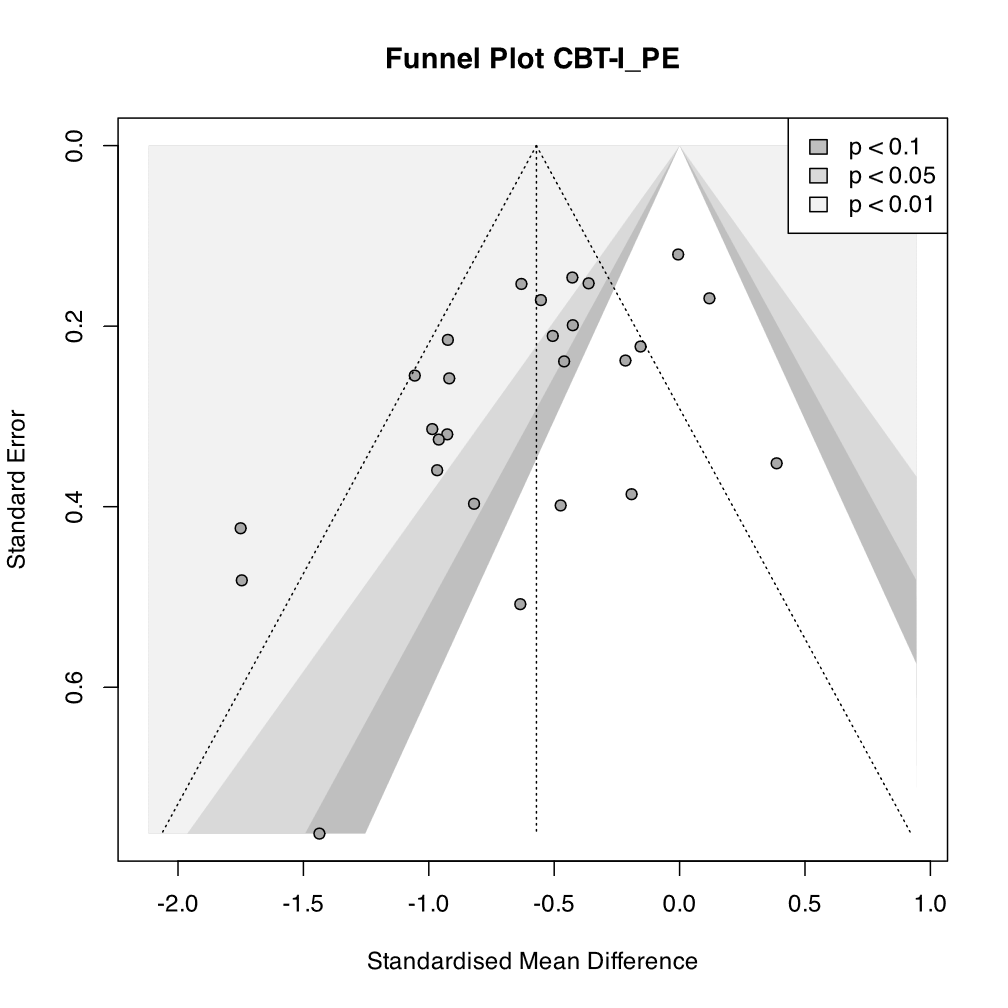

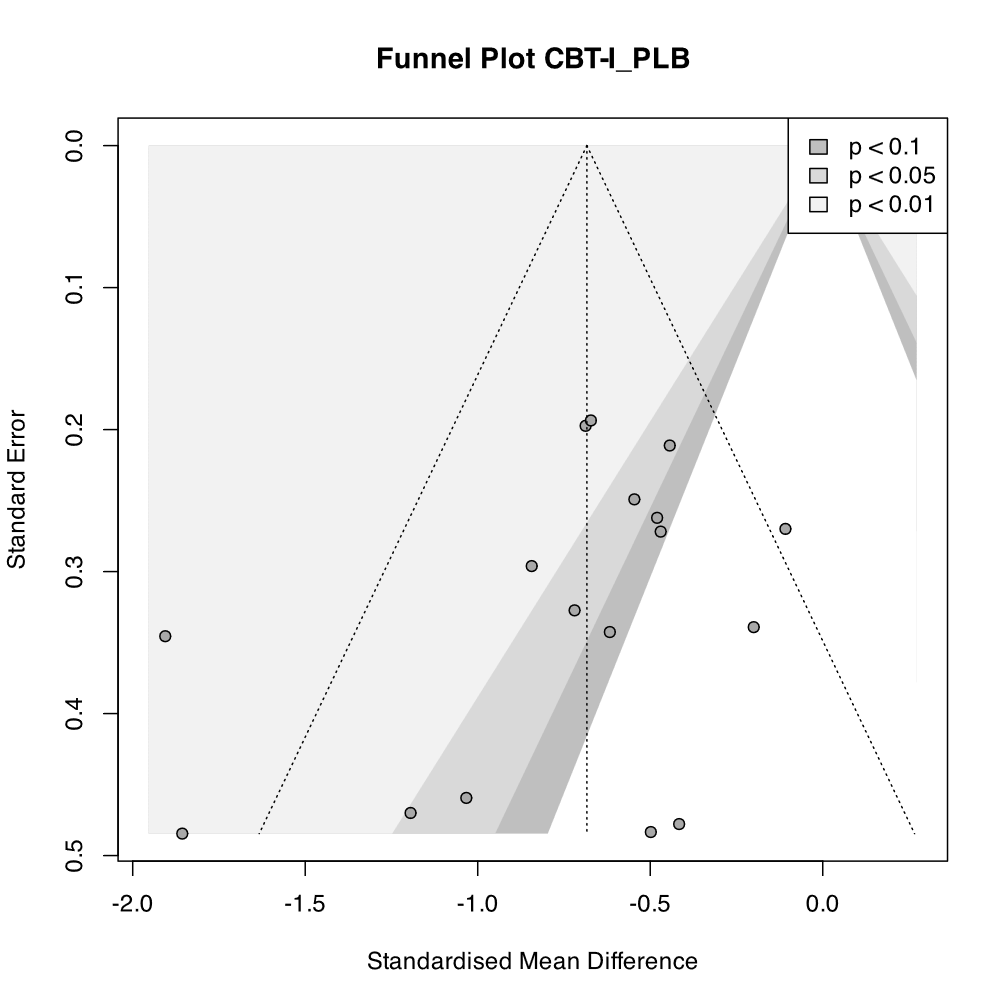

### 4. PRIMARY OUTCOME (Insomnia remission)

#### Direct estimate and indirect estimates of the network meta-anlaysis

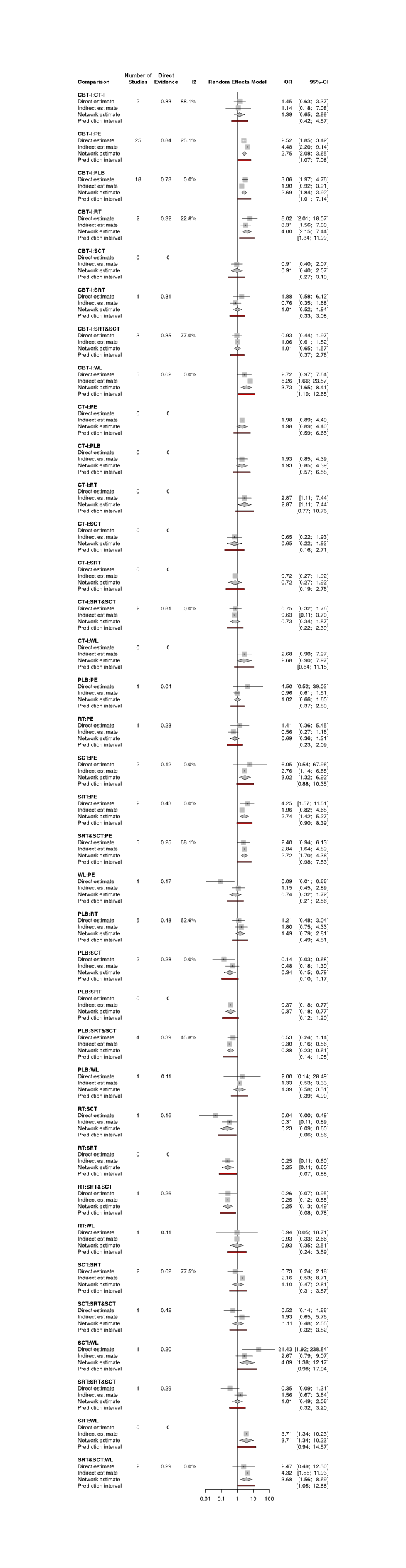

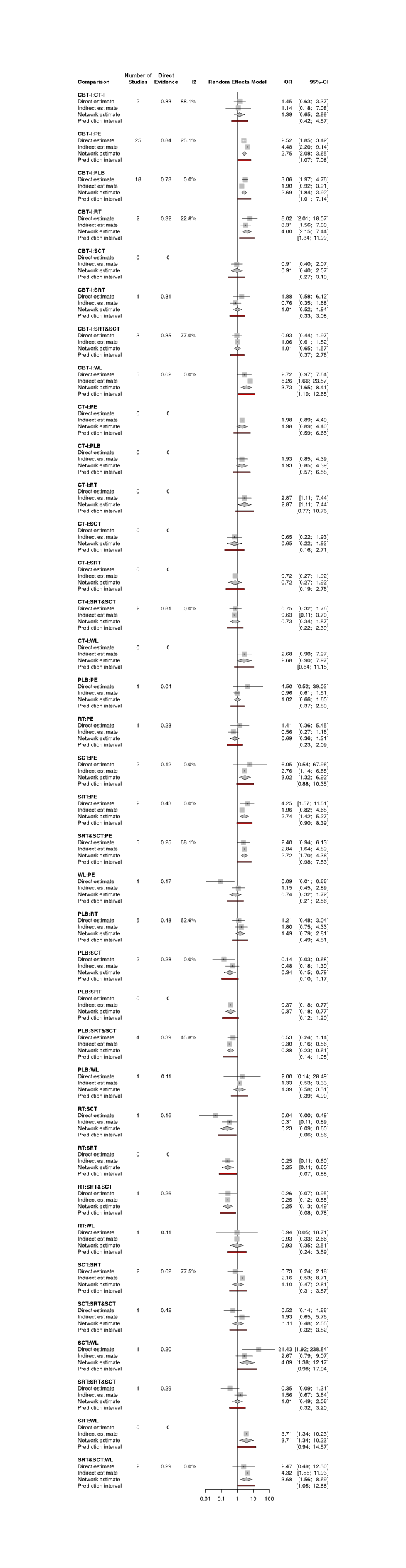

### 5. CONFIDENCE IN NETWORK META-ANALYSIS (CINeMA)

We used the average RoB for the within-study bias domain. We upgraded the within-study bias domain for CBT-I:PE, CBT-I:RT, PE:SCT, and PE:SRT&SCTbased on sensitivity analyses excluding high risk of bias and some concerns trials. For the reporting bias domain, we set “some concerns” as default, because the lack of preregistration is still prevalent in the field, and visual inspection of funnel plots suggested some reporting bias. However, we did not downgrade for this domain for CBT-I:PE and CBT-I:PLB because the results were constantly OR > 1, and we judged adjusting for publication bias would not change the result. We upgraded for those comparing active interventions (CBT-I, SRT&SCT, SRT, SCT, CT-I and RT). We set the clinically important size of effect as OR = 1.2. We downgraded the confidence rating by two levels for each domain with major concerns, and by one level for each domain with some concerns. Given the interactions between the imprecision, heterogeneity and incoherence domains, we did not downgrade twice for these domains.

| **Comparison** | **k** | **Within-study bias** | **Reporting bias** | **Indirectness** | **Imprecision** | **Heterogeneity** | **Incoherence** | **Confidence rating** |
| --- | --- | --- | --- | --- | --- | --- | --- | --- |
| CBT-I:CT-I | 2 | No concerns | No concerns | No concerns | Major concerns | No concerns | No concerns | Low |
| CBT-I:PE | 25 | No concerns | Some concerns | No concerns | No concerns | Some concerns | No concerns | Moderate |
| CBT-I:PLB | 18 | Some concerns | Some concerns | No concerns | No concerns | Some concerns | No concerns | Low |
| CBT-I:RT | 2 | No concerns | No concerns | No concerns | No concerns | No concerns | No concerns | High |
| CBT-I:SRT | 1 | Some concerns | No concerns | No concerns | Major concerns | No concerns | No concerns | Very low |
| CBT-I:SRT&SCT | 3 | Some concerns | No concerns | No concerns | Major concerns | No concerns | No concerns | Very low |
| CBT-I:WL | 5 | Some concerns | Some concerns | No concerns | No concerns | No concerns | No concerns | Low |
| CT-I:SRT&SCT | 2 | No concerns | No concerns | No concerns | Major concerns | No concerns | No concerns | Low |
| PE:PLB | 1 | Some concerns | Some concerns | No concerns | Major concerns | No concerns | No concerns | Very low |
| PE:RT | 1 | Some concerns | Some concerns | No concerns | Major concerns | No concerns | No concerns | Very low |
| PE:SCT | 2 | No concerns | Some concerns | No concerns | No concerns | Major concerns | No concerns | Very low |
| PE:SRT | 2 | Some concerns | Some concerns | No concerns | No concerns | Major concerns | No concerns | Very low |
| PE:SRT&SCT | 5 | No concerns | Some concerns | No concerns | No concerns | Some concerns | No concerns | Low |
| PE:WL | 1 | Some concerns | Some concerns | No concerns | Major concerns | No concerns | Major concerns | Very low |
| PLB:RT | 5 | Some concerns | Some concerns | No concerns | Major concerns | No concerns | No concerns | Very low |
| PLB:SCT | 2 | Some concerns | Some concerns | No concerns | No concerns | Major concerns | No concerns | Very low |
| PLB:SRT&SCT | 4 | Some concerns | Some concerns | No concerns | No concerns | Some concerns | No concerns | Very low |
| PLB:WL | 1 | Some concerns | Some concerns | No concerns | Major concerns | No concerns | No concerns | Very low |
| RT:SCT | 2 | Some concerns | No concerns | No concerns | No concerns | Some concerns | No concerns | Very low |
| RT:SRT&SCT | 1 | Some concerns | No concerns | No concerns | No concerns | No concerns | No concerns | Moderate |
| RT:WL | 1 | Some concerns | Some concerns | No concerns | Major concerns | No concerns | No concerns | Very low |
| SCT:SRT | 2 | Some concerns | No concerns | No concerns | Major concerns | No concerns | No concerns | Very low |
| SCT:SRT&SCT | 1 | Some concerns | No concerns | No concerns | Major concerns | No concerns | No concerns | Very low |
| SCT:WL | 1 | Some concerns | Some concerns | No concerns | No concerns | Some concerns | No concerns | Very low |
| SRT:SRT&SCT | 1 | Some concerns | No concerns | No concerns | Major concerns | No concerns | No concerns | Very low |
| SRT&SCT:WL | 2 | Some concerns | Some concerns | No concerns | No concerns | No concerns | No concerns | Low |
| CBT-I:SCT | 0 | Some concerns | No concerns | No concerns | Major concerns | No concerns | No concerns | Very low |
| CT-I:PE | 0 | Some concerns | Some concerns | No concerns | Some concerns | Some concerns | No concerns | Very low |
| CT-I:PLB | 0 | Some concerns | Some concerns | No concerns | Some concerns | Some concerns | No concerns | Very low |
| CT-I:RT | 0 | Some concerns | No concerns | No concerns | Some concerns | Some concerns | No concerns | Very low |
| CT-I:SCT | 0 | Some concerns | No concerns | No concerns | Major concerns | No concerns | No concerns | Very low |
| CT-I:SRT | 0 | Some concerns | No concerns | No concerns | Major concerns | No concerns | No concerns | Very low |
| CT-I:WL | 0 | Some concerns | Some concerns | No concerns | Some concerns | Some concerns | No concerns | Very low |
| PLB:SRT | 0 | Some concerns | Some concerns | No concerns | No concerns | Major concerns | No concerns | Very low |
| RT:SRT | 0 | Some concerns | No concerns | No concerns | No concerns | Some concerns | No concerns | Low |
| SRT:WL | 0 | Some concerns | Some concerns | No concerns | No concerns | Some concerns | No concerns | Very low |

### 6. SECONDARY OUTCOMES

#### (i) Dropout vs PE (dichotomous)

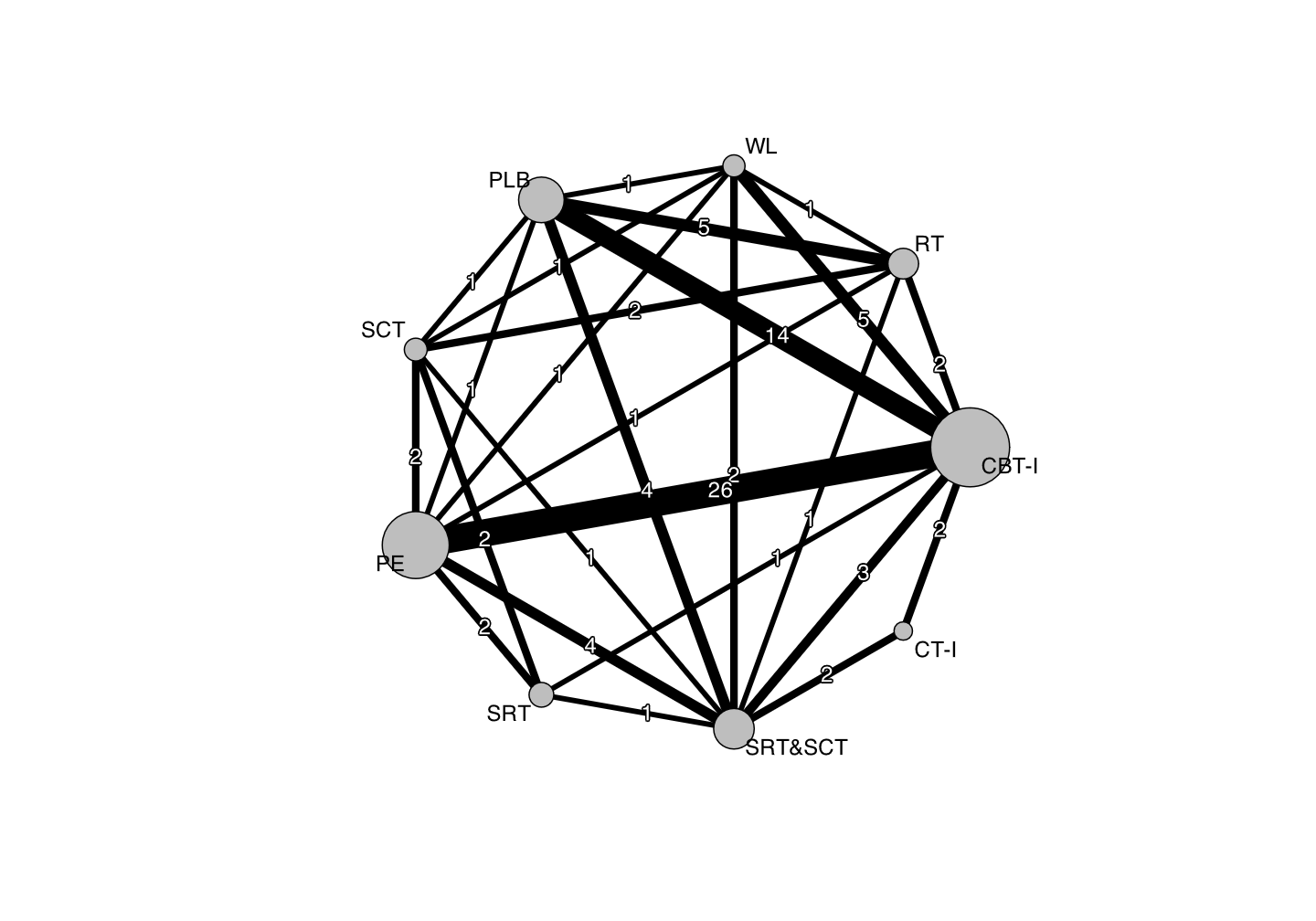

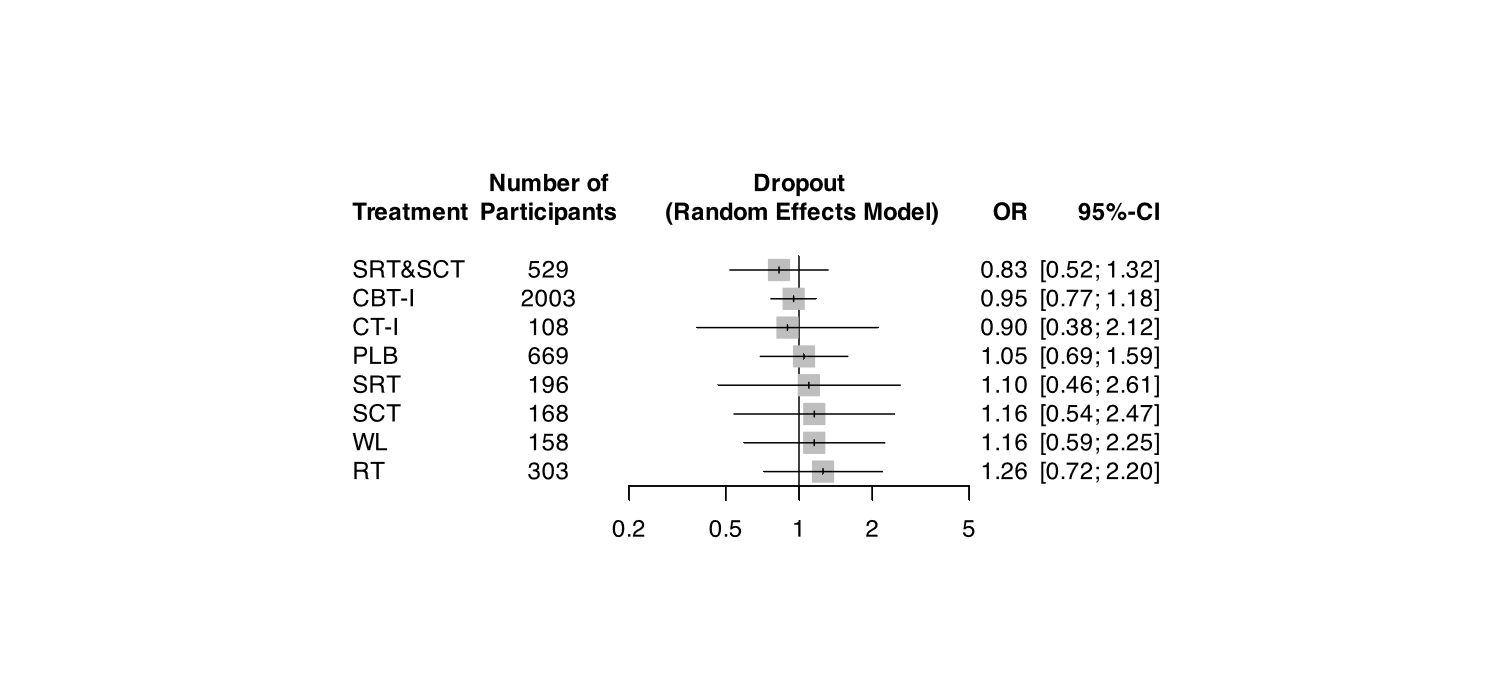

#### (ii) Sleep efficiency vs PE (%, continuous)

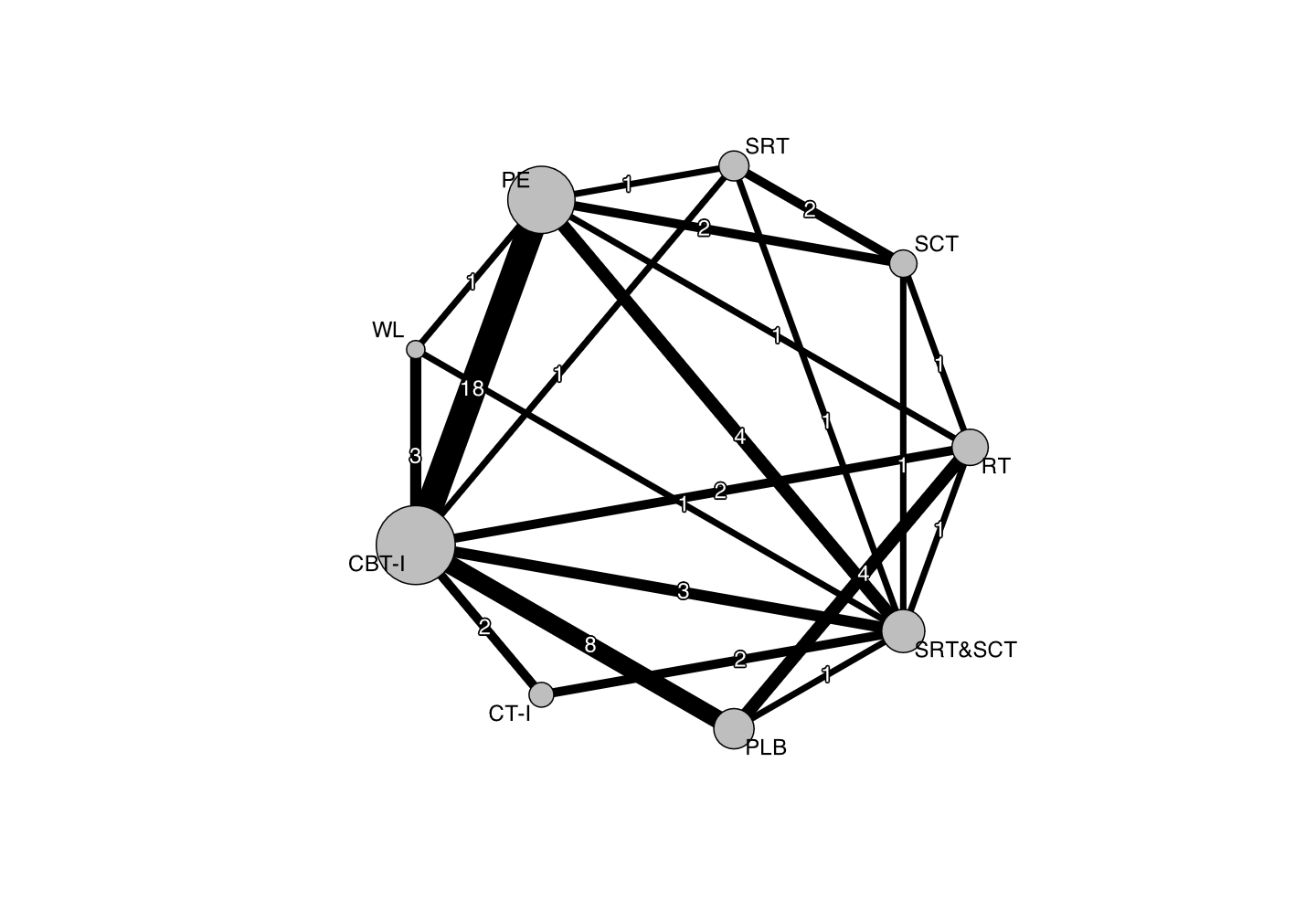

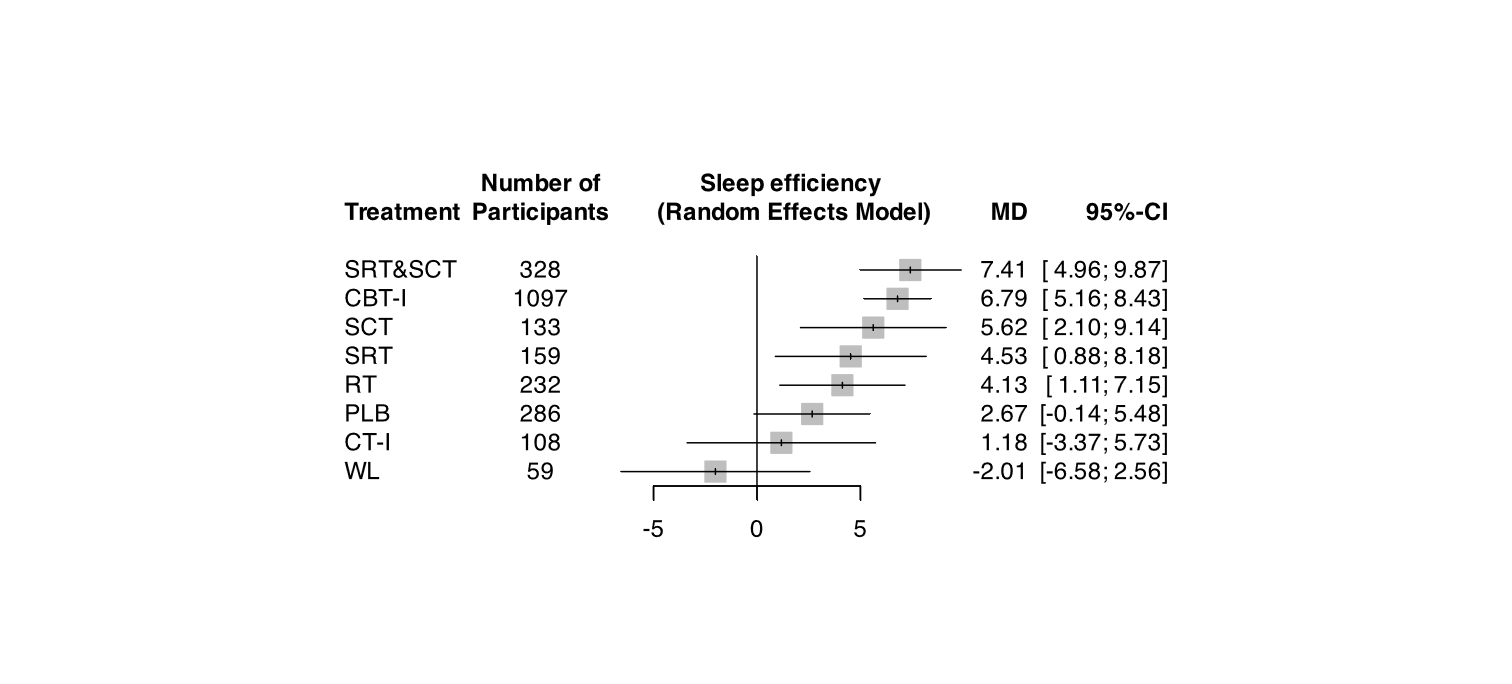

#### (iii) Sleep latency vs PE (minutes, continuous)

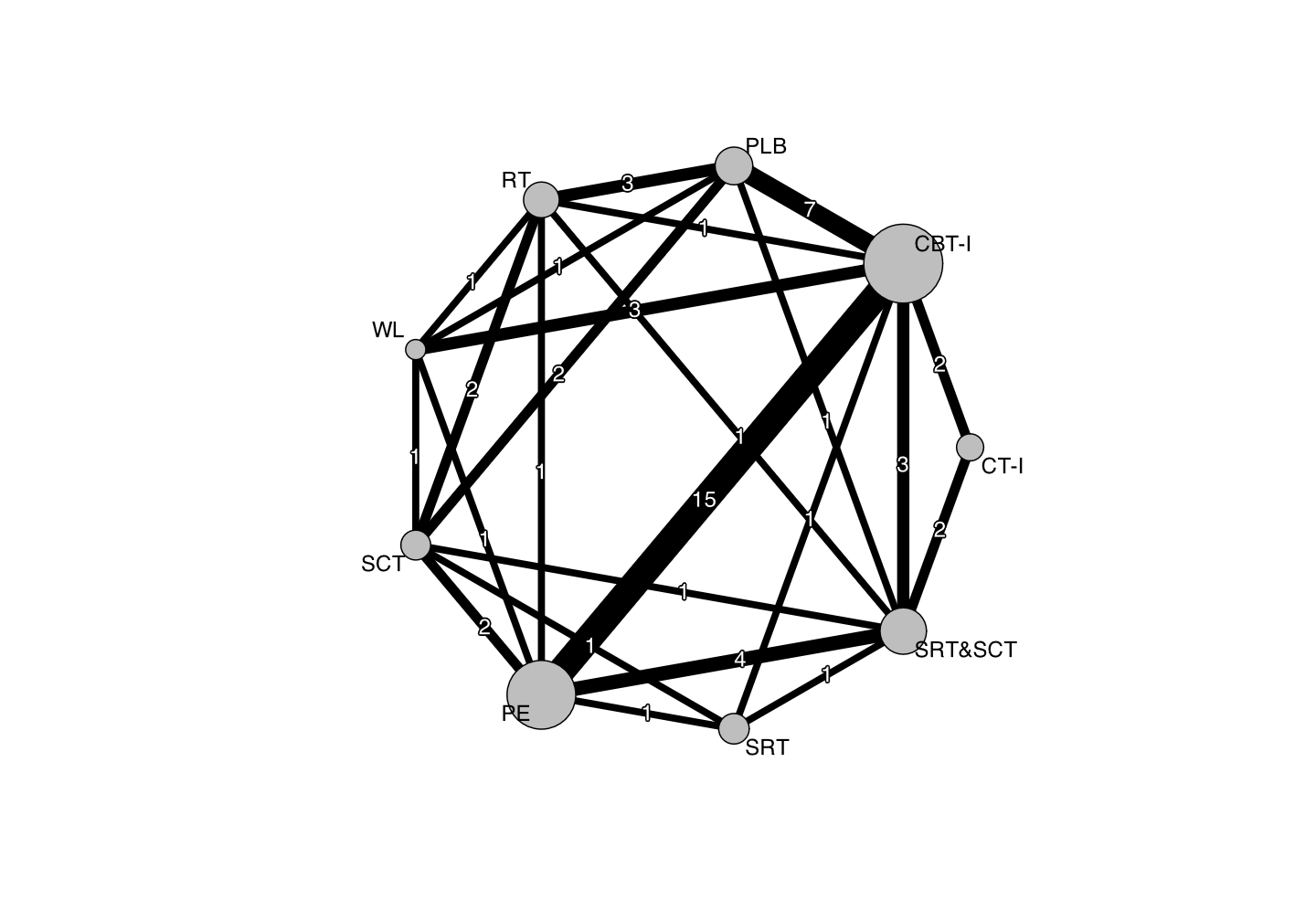

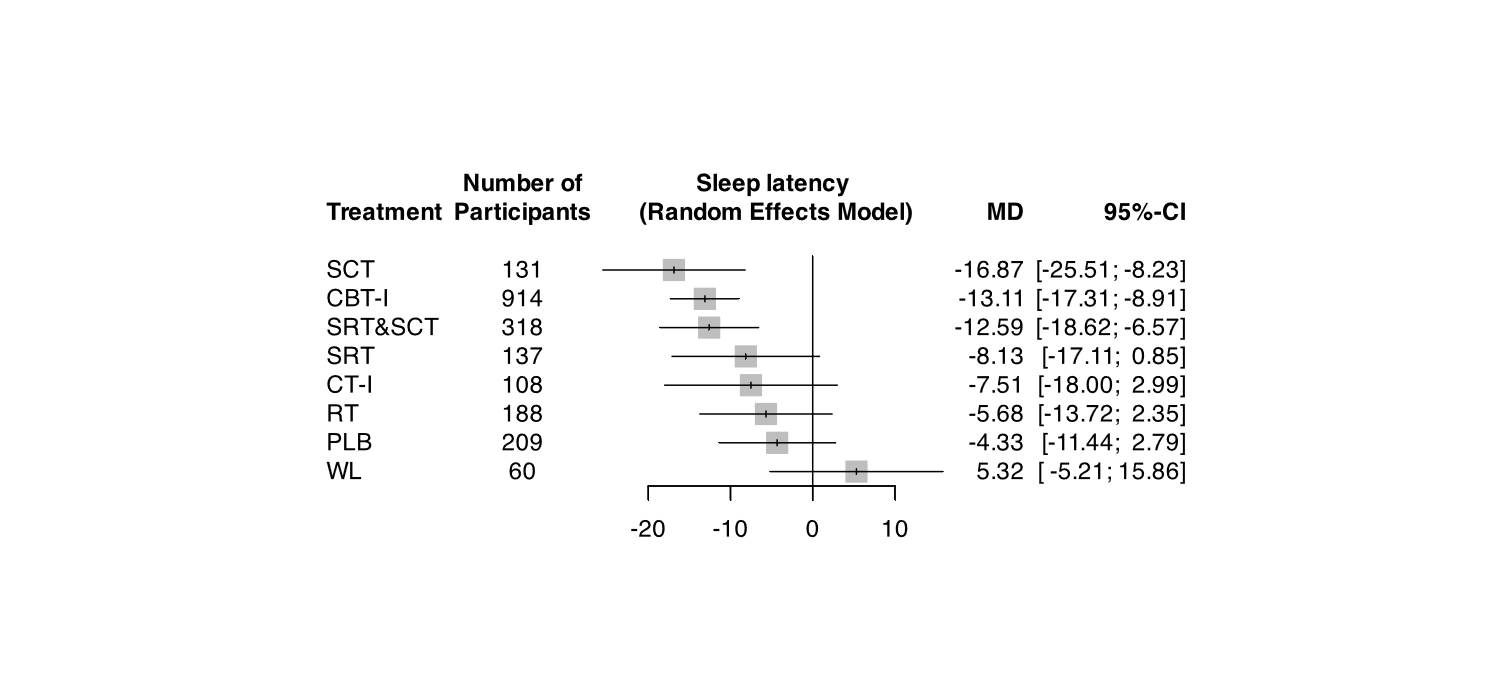

#### (iv) Wake after sleep onset (minutes, continuous)

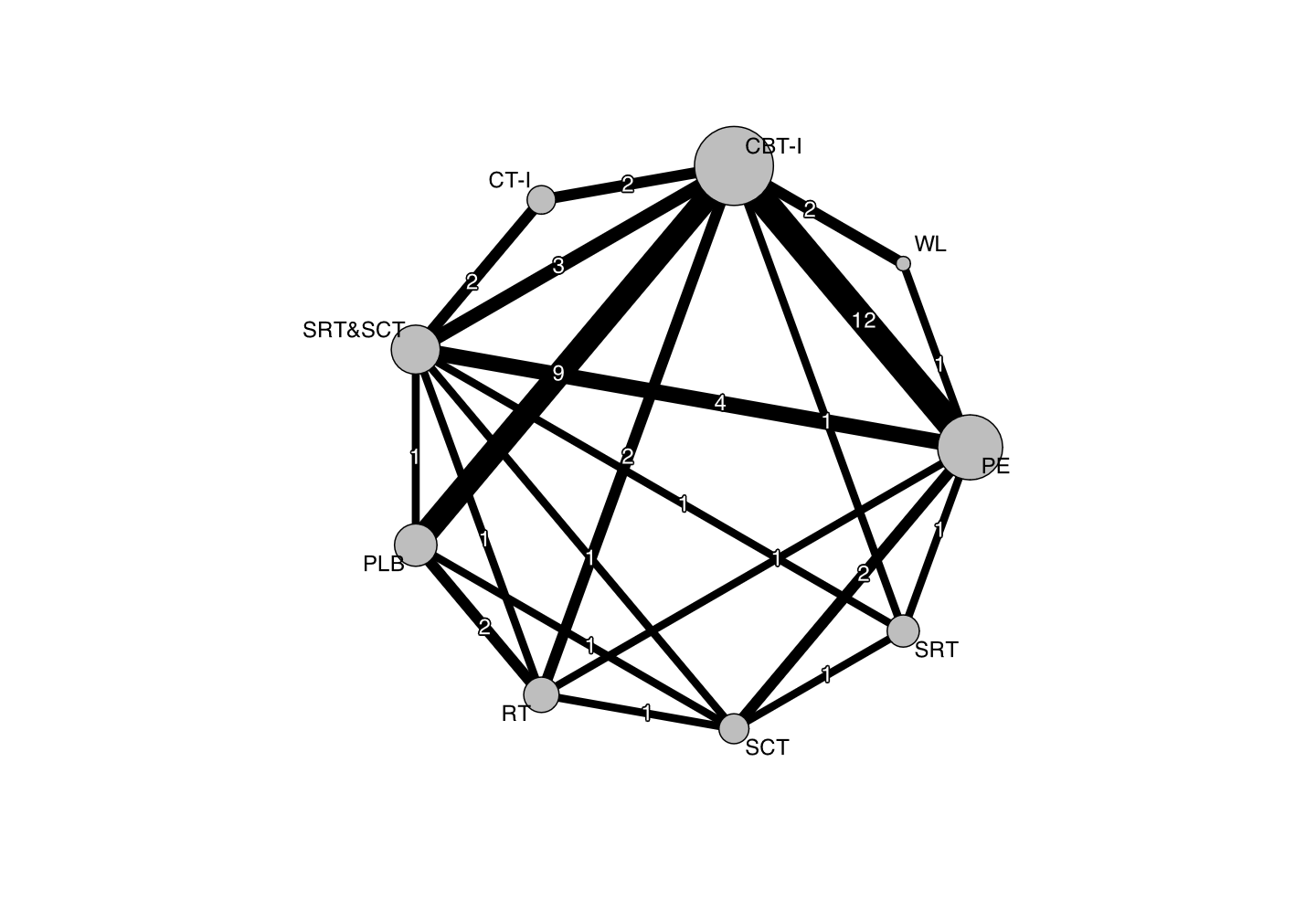

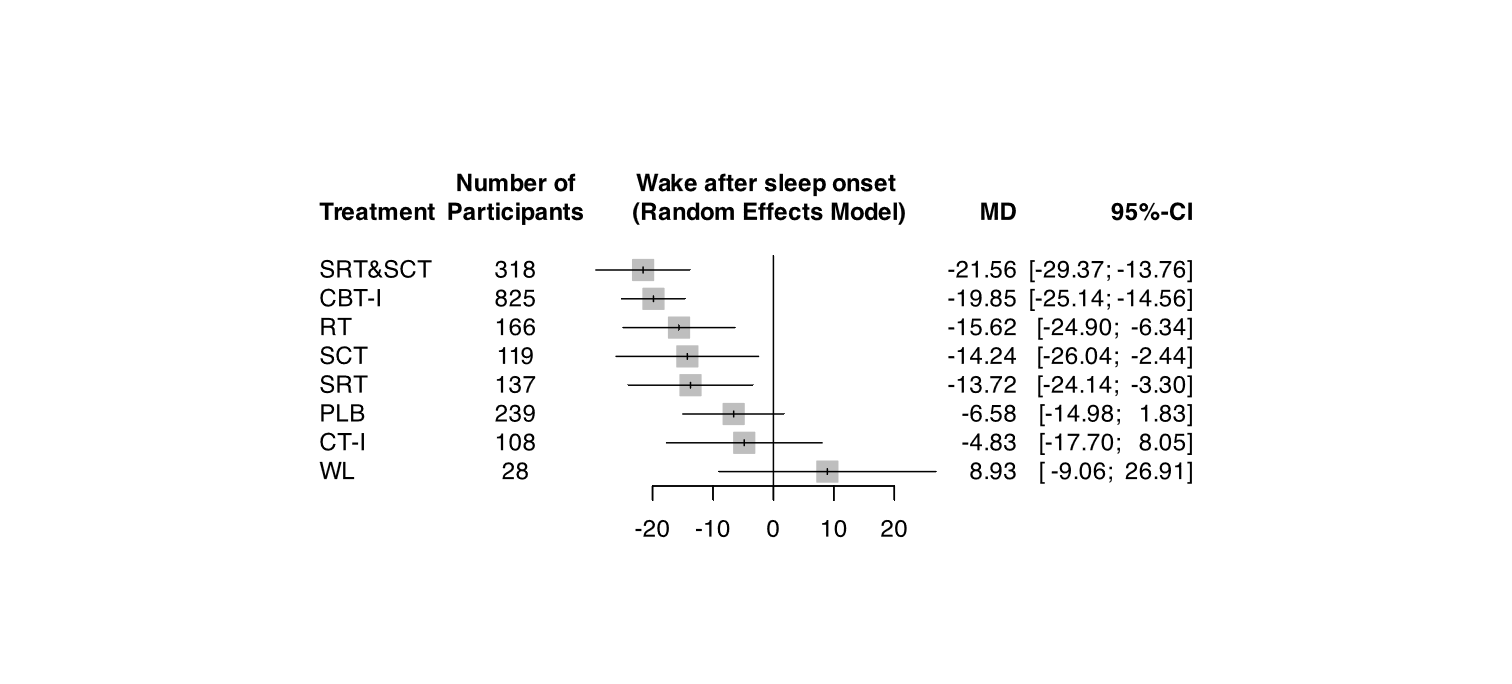

#### (v) Total sleep time (minutes, continuous)

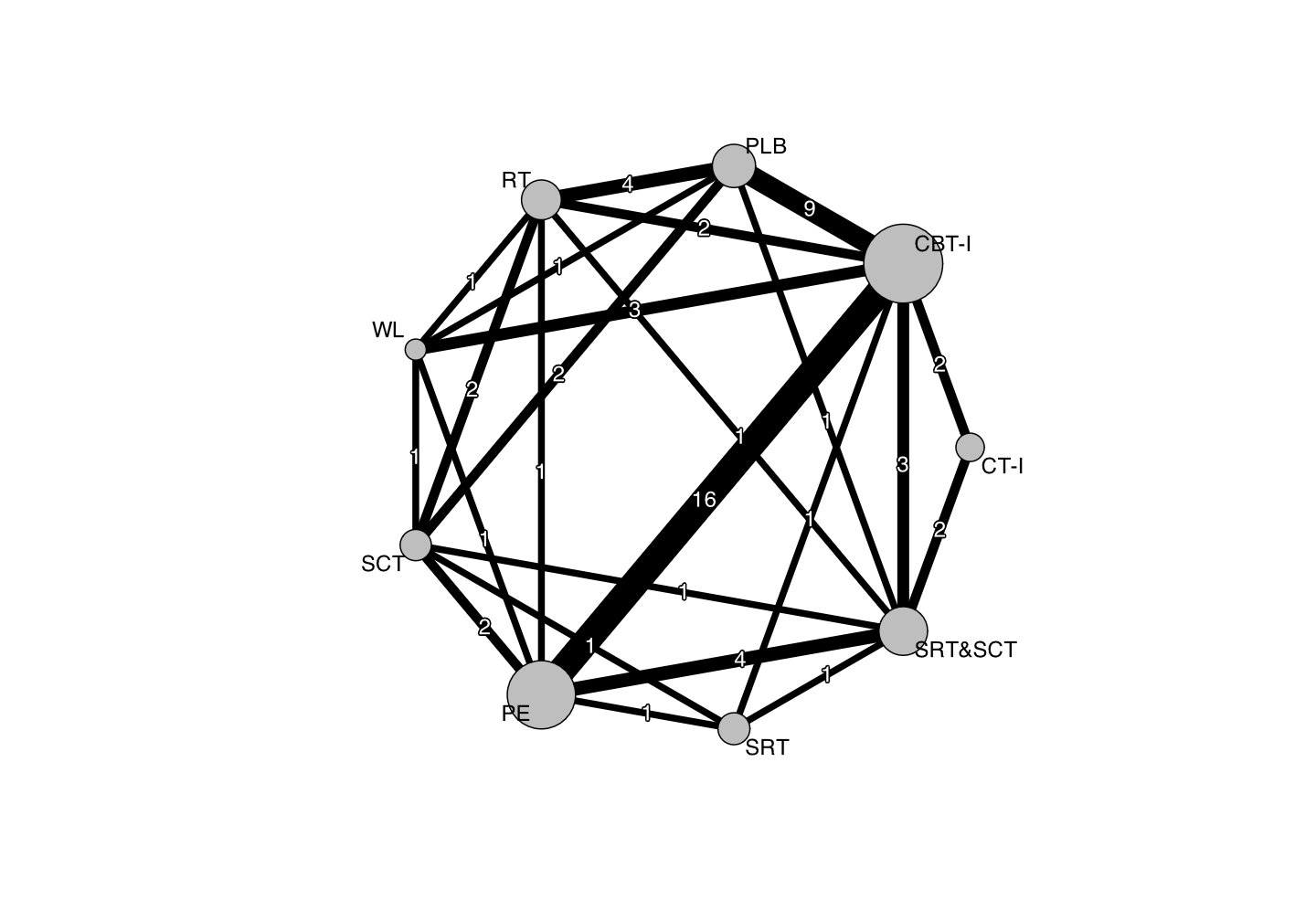

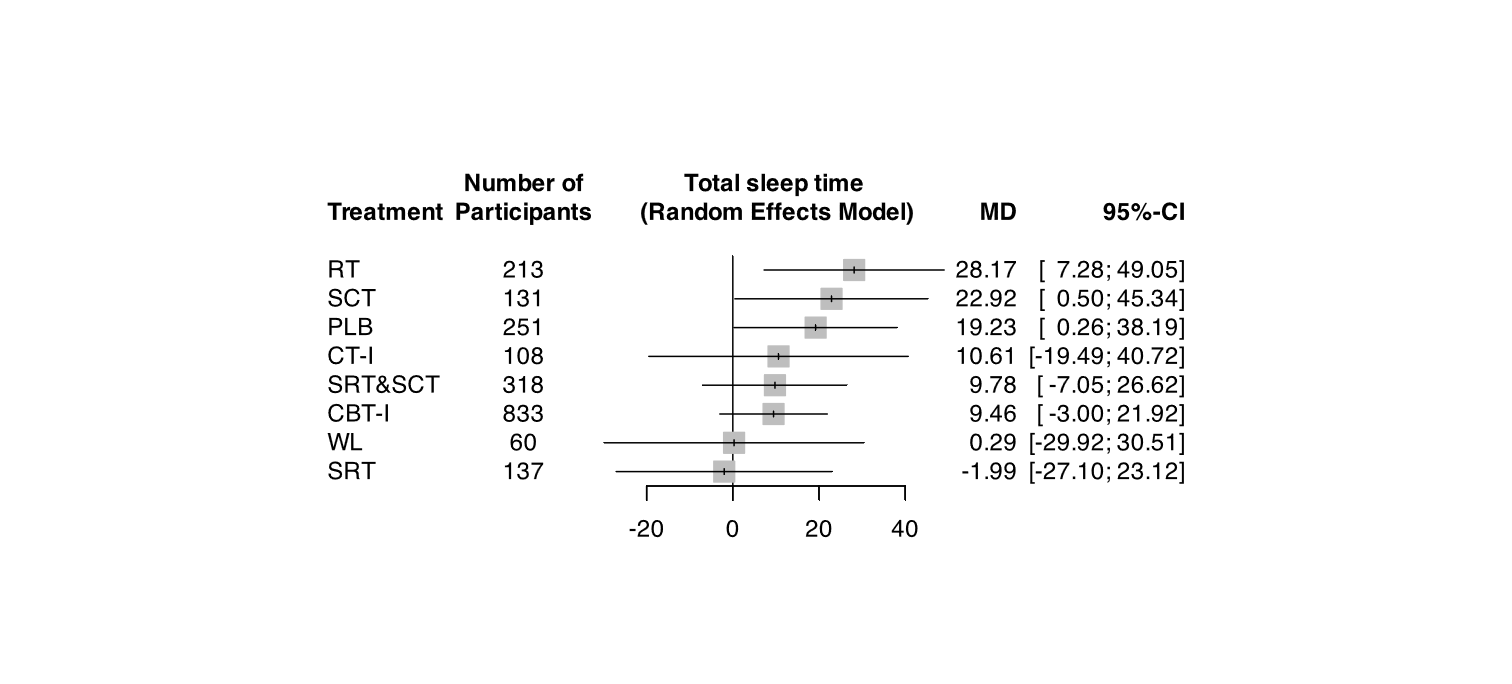

#### (vi) Insomnia severity (continuous)

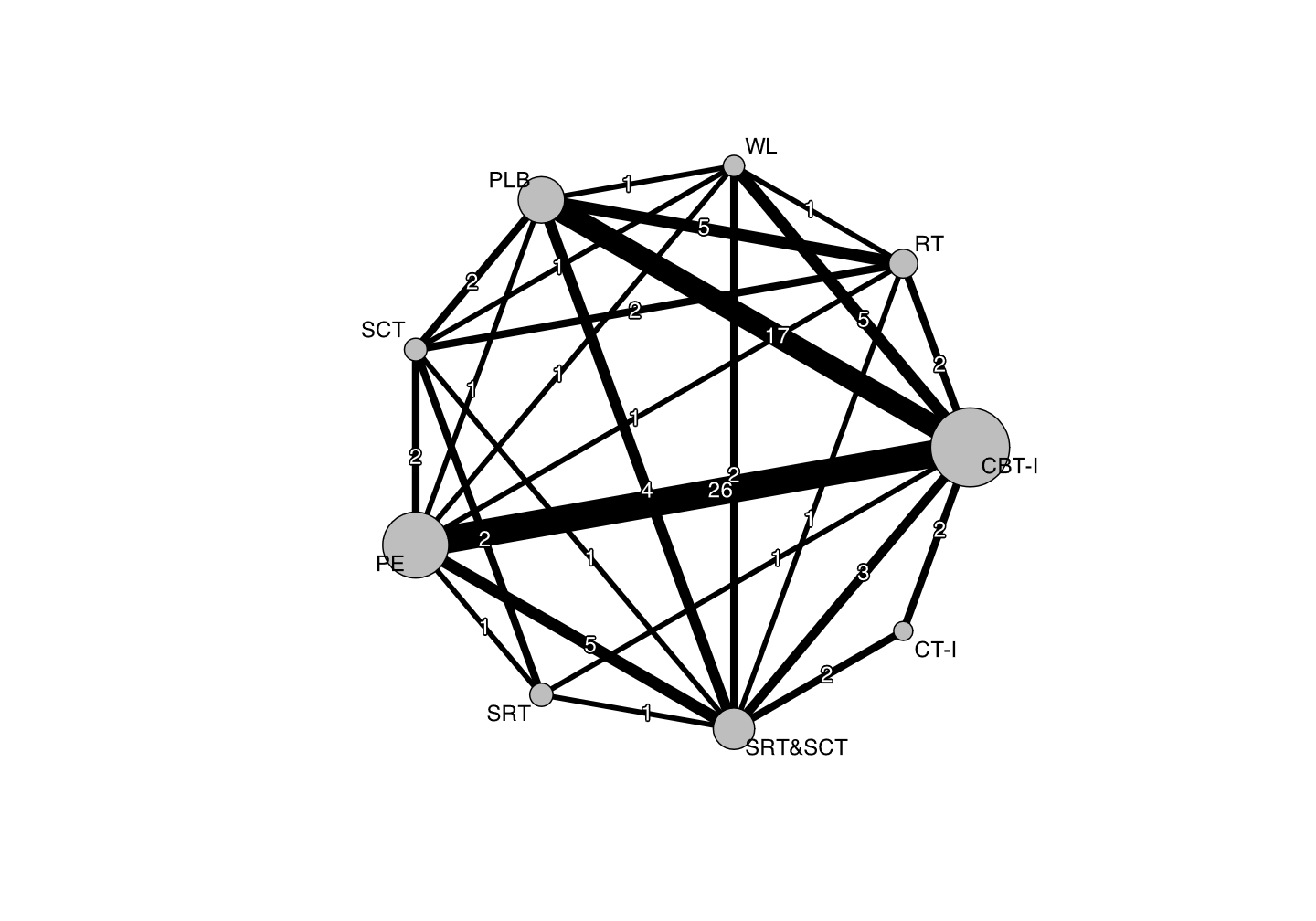

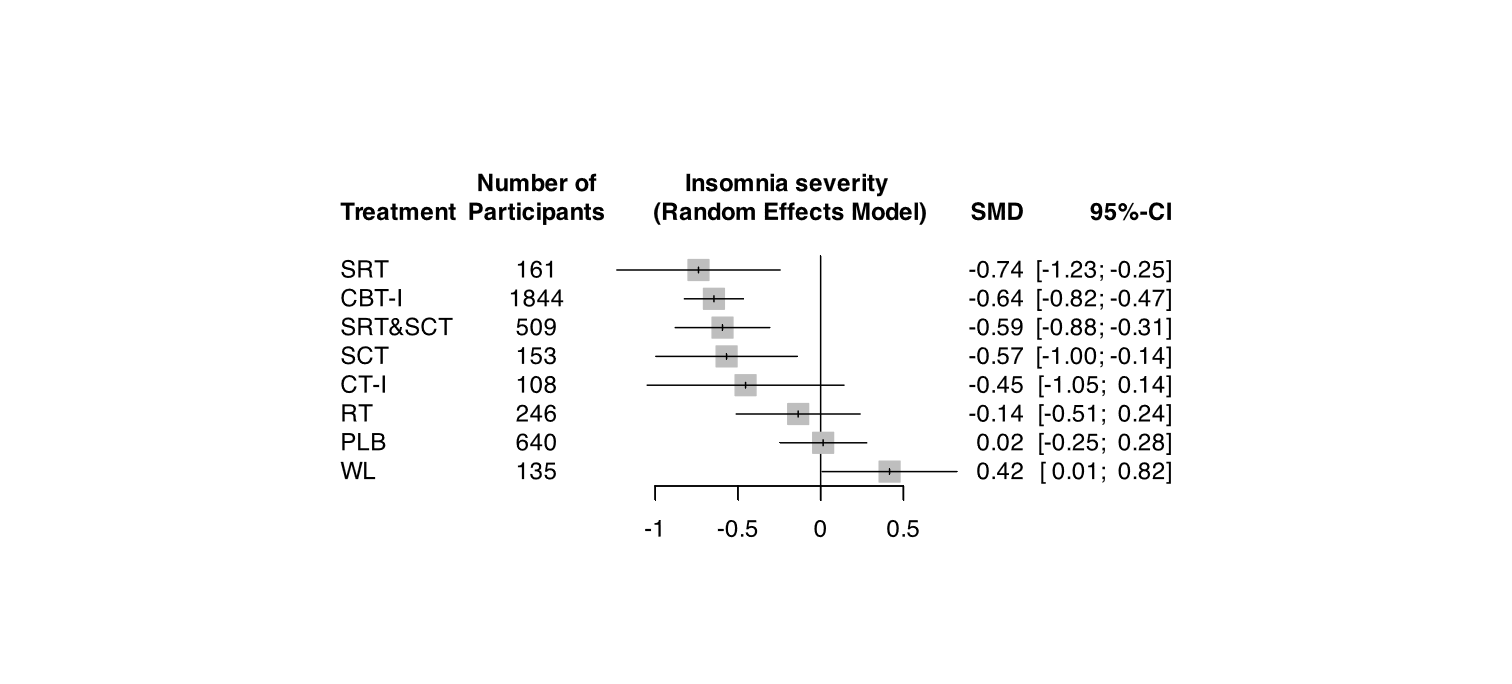

#### (vii) Insomnia remission, long-term (dichotomous)

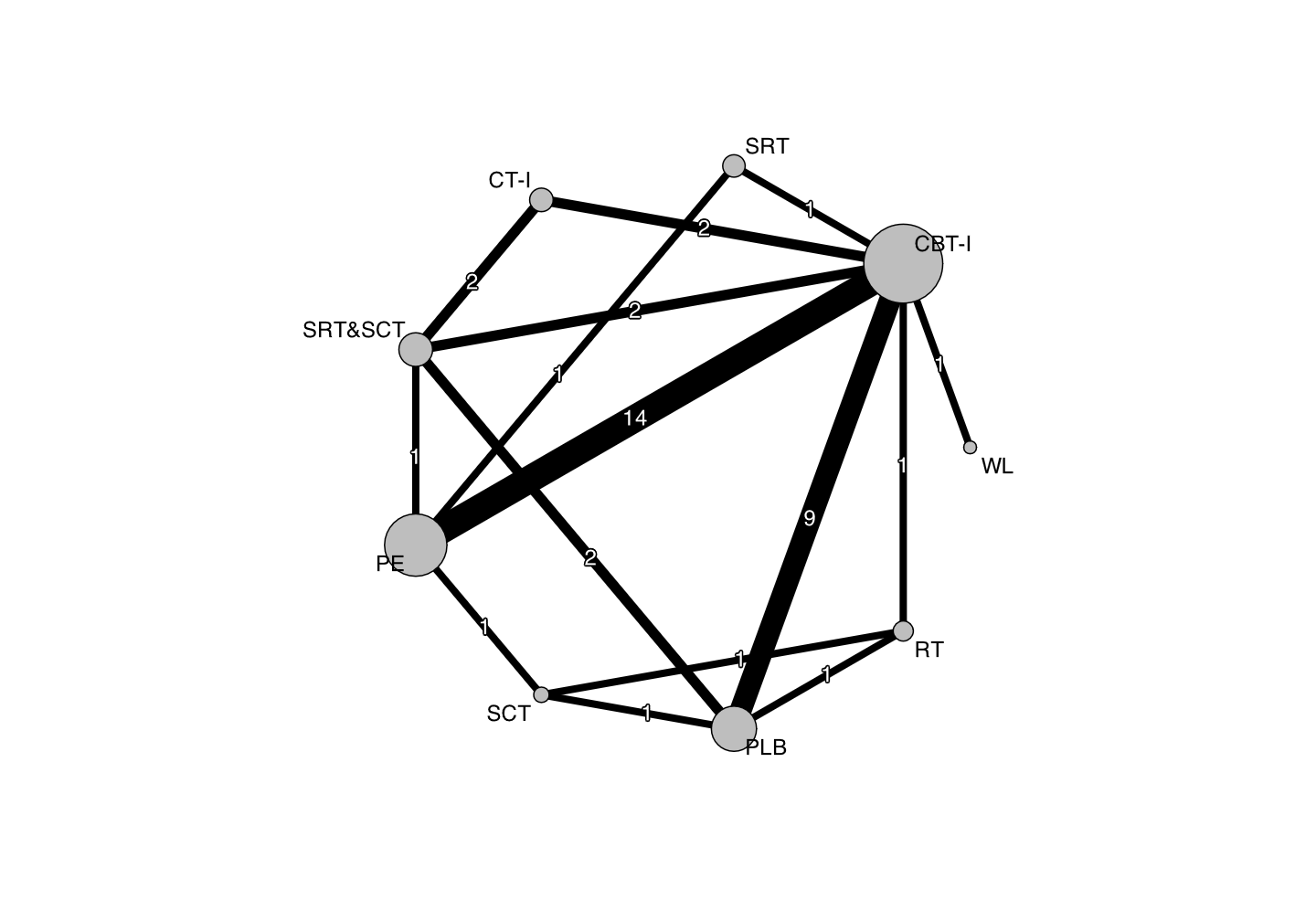

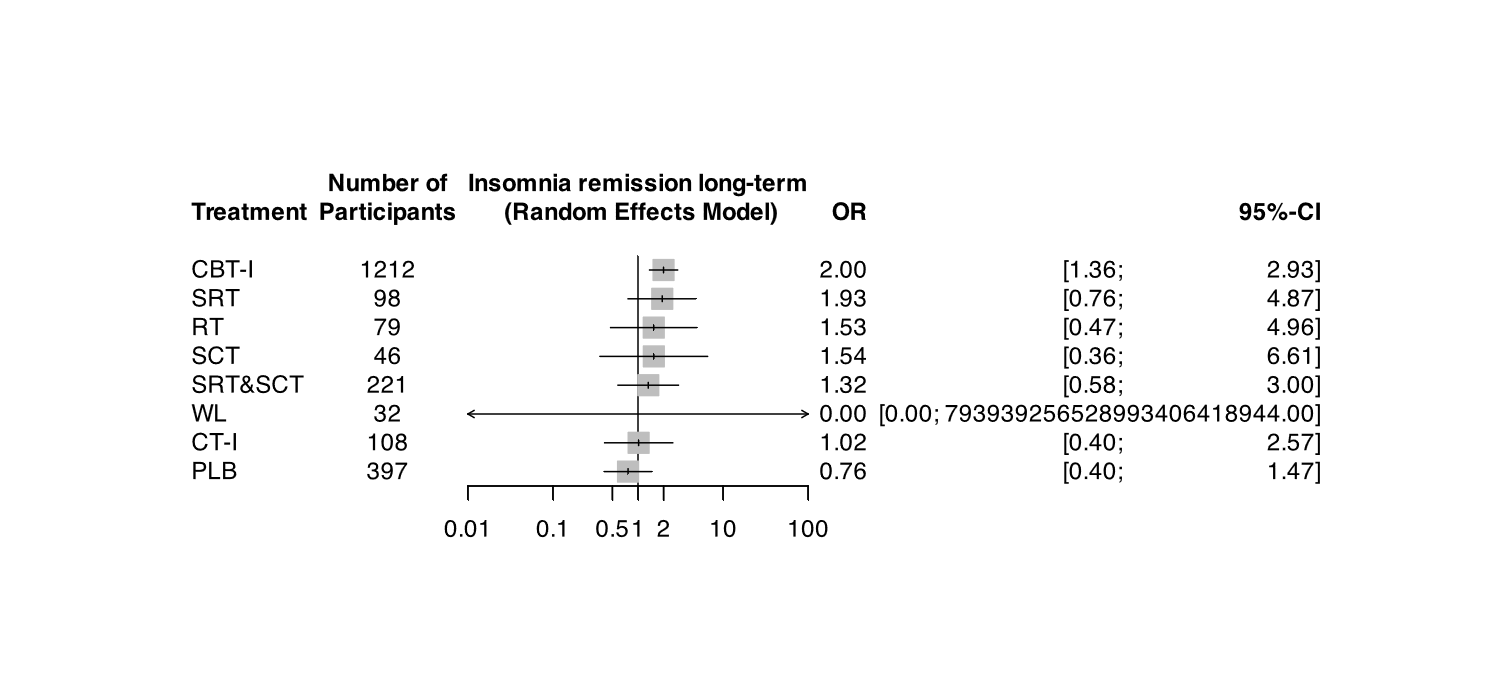

#### (viii) Insomnia severity, long-term (continuous)

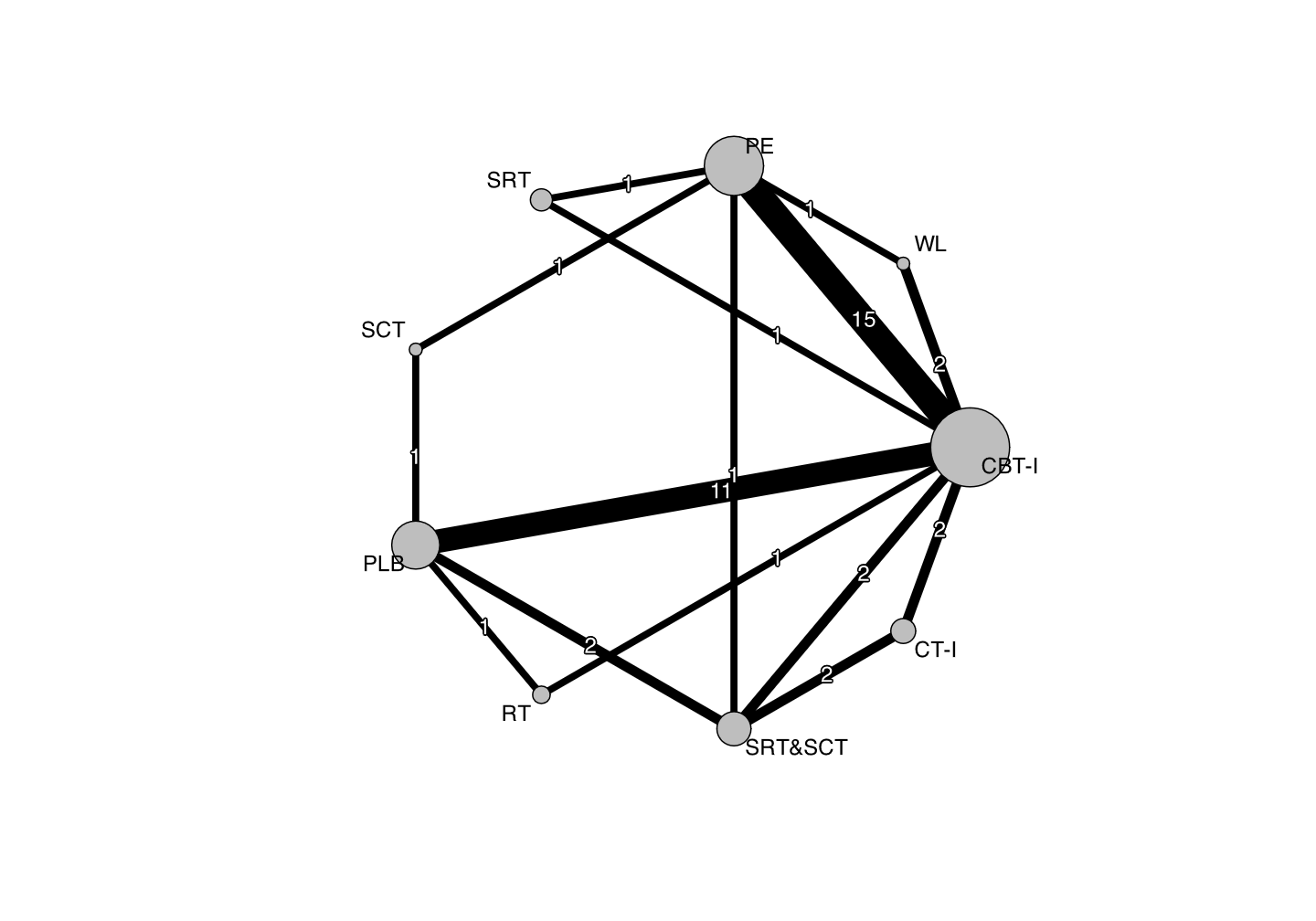

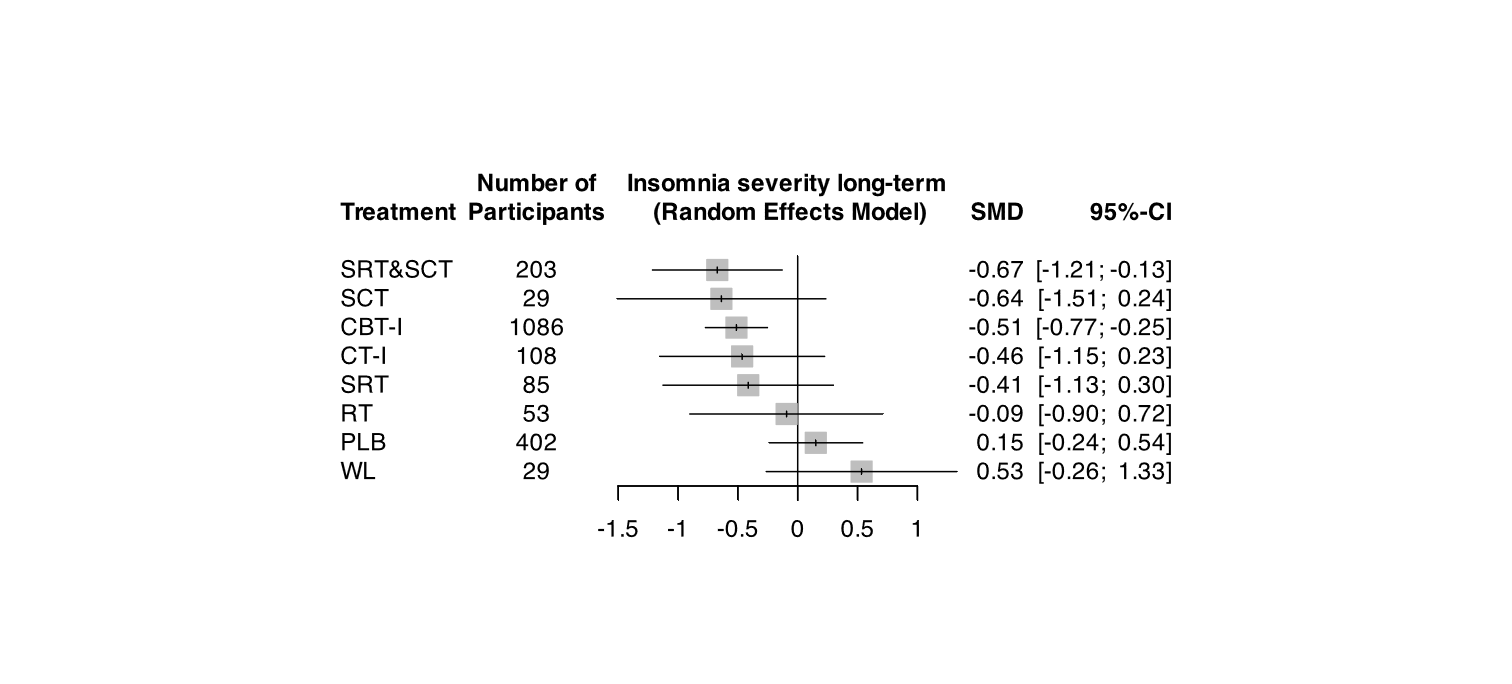

### 7. SENSITIVITY ANALYSES

#### S1. Formal diagnosis only

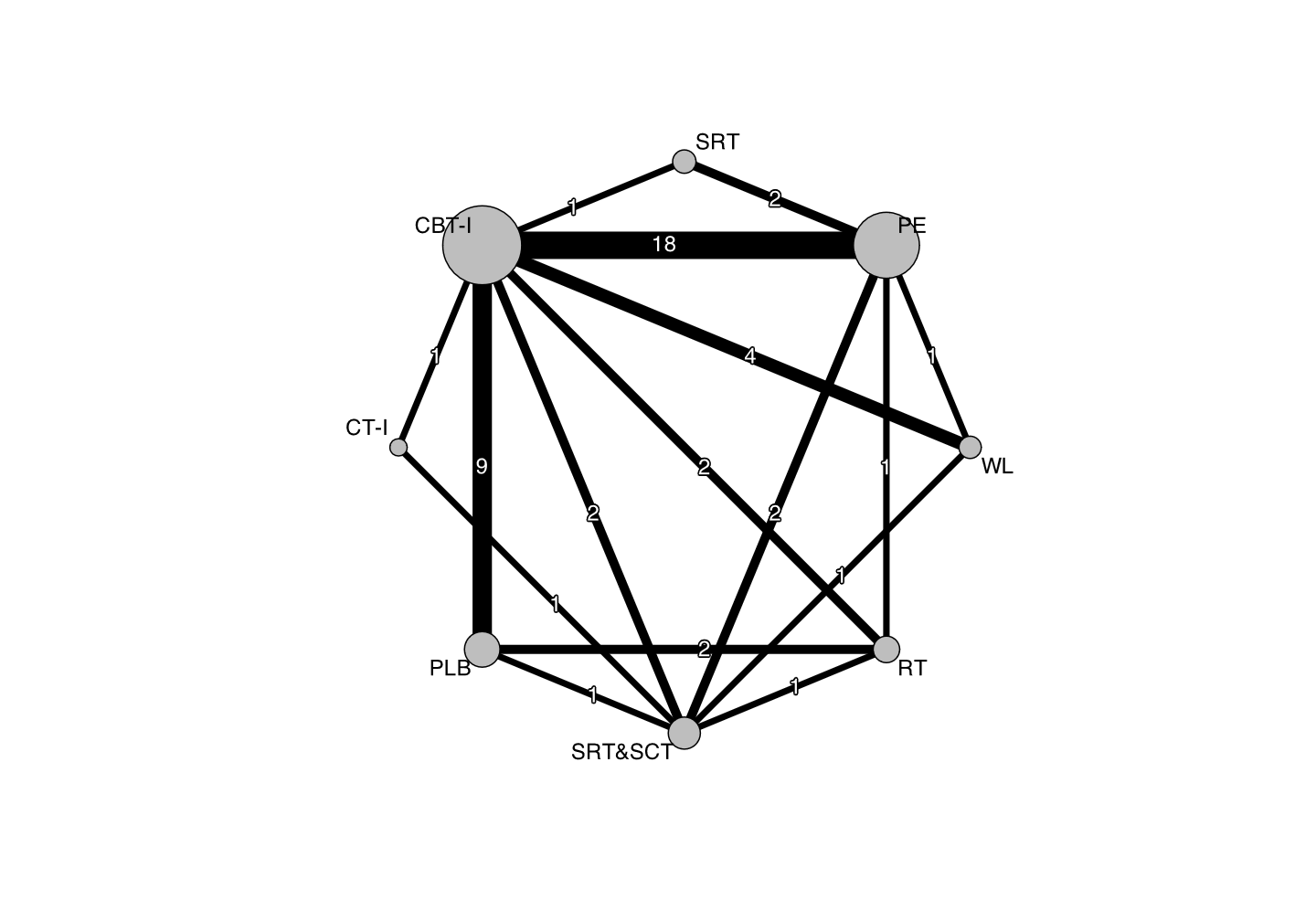

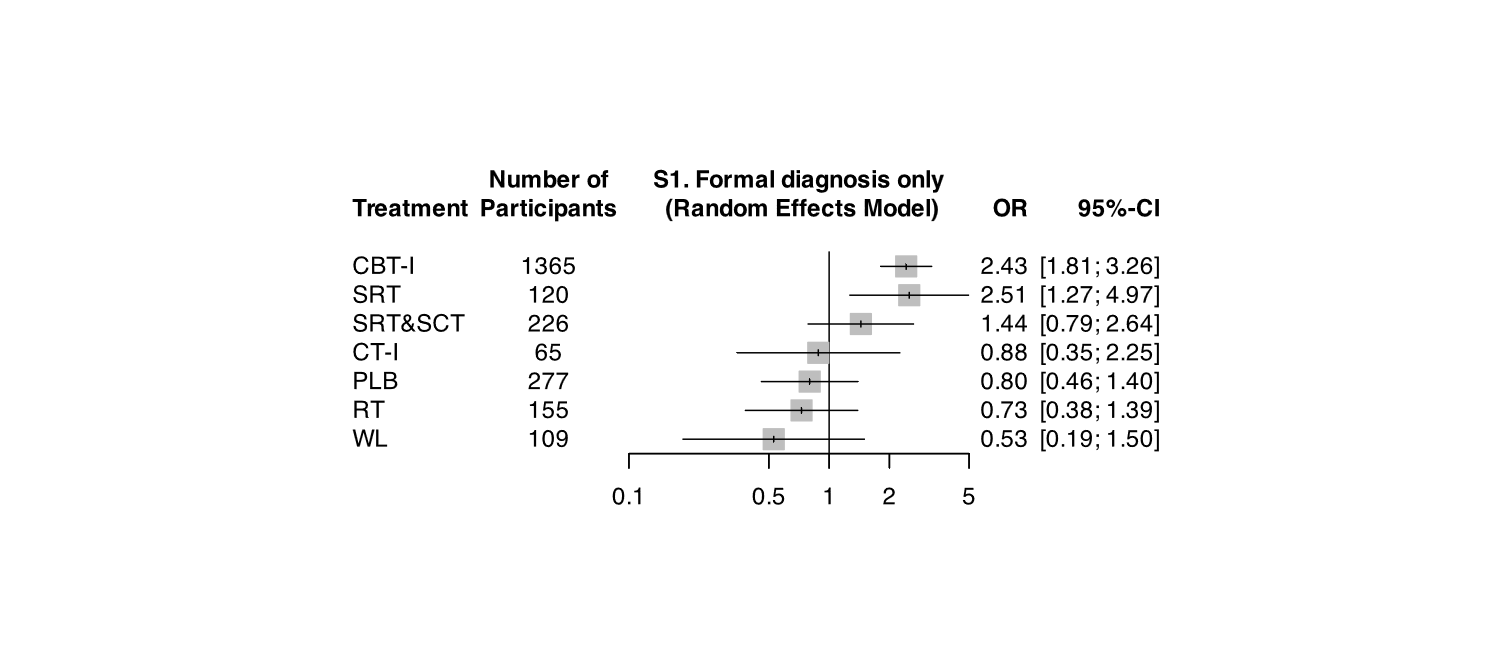

#### S2. Insomnia without comorbidity only

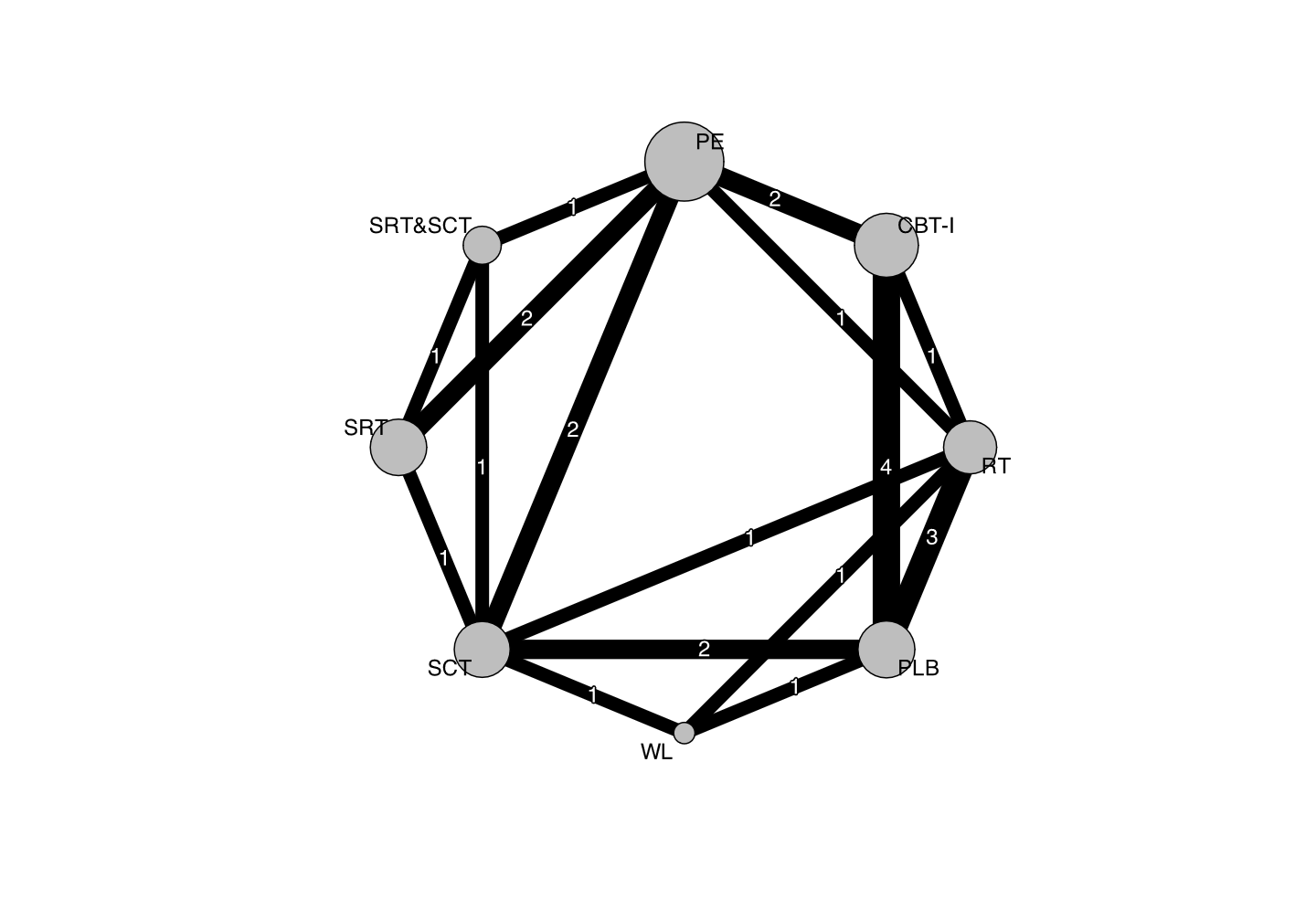

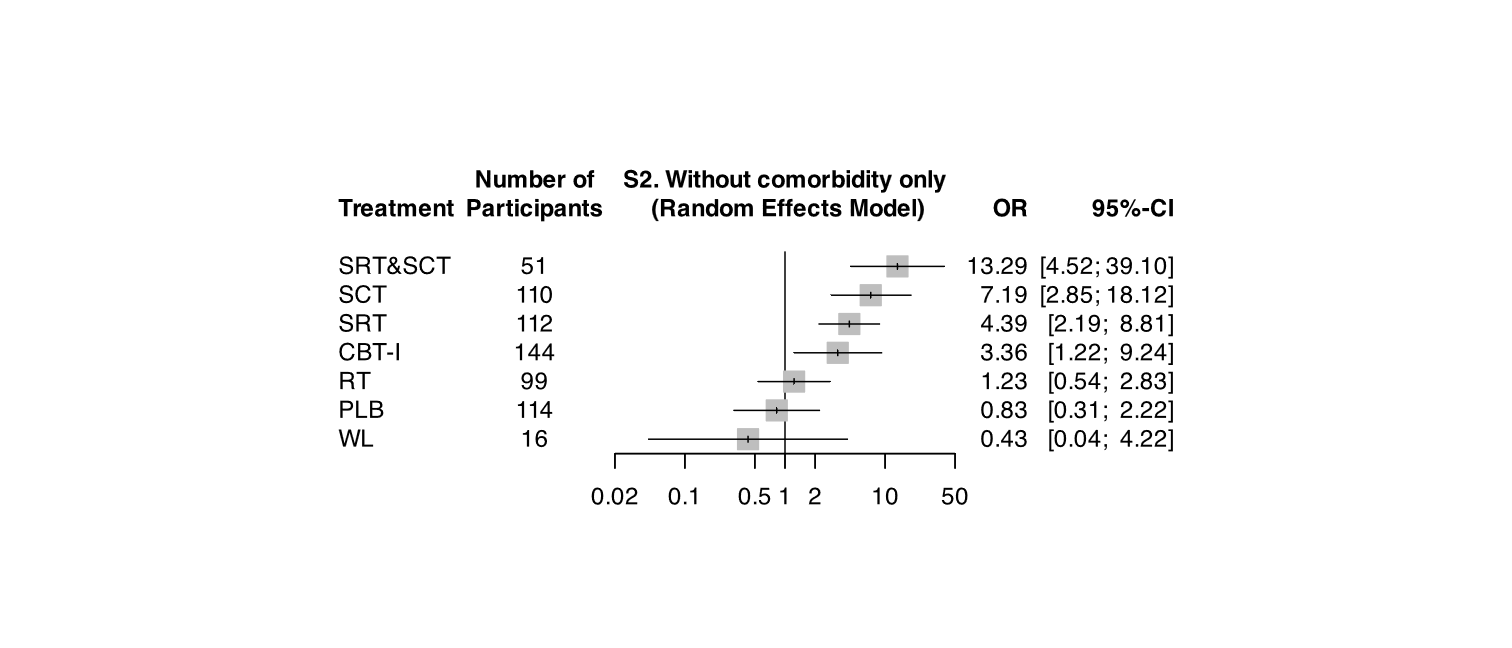

#### S3. Excluding trials with high dropout rate

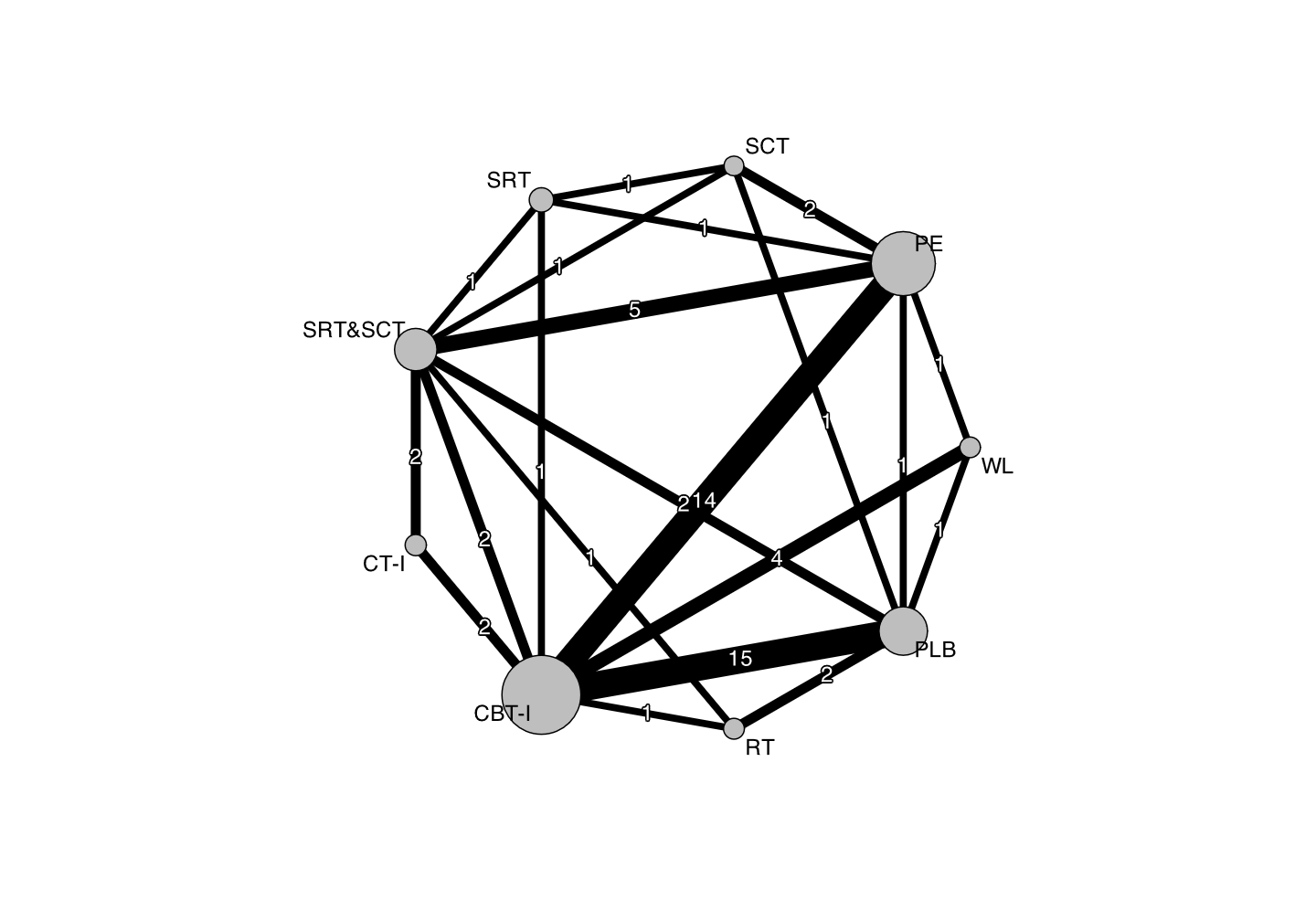

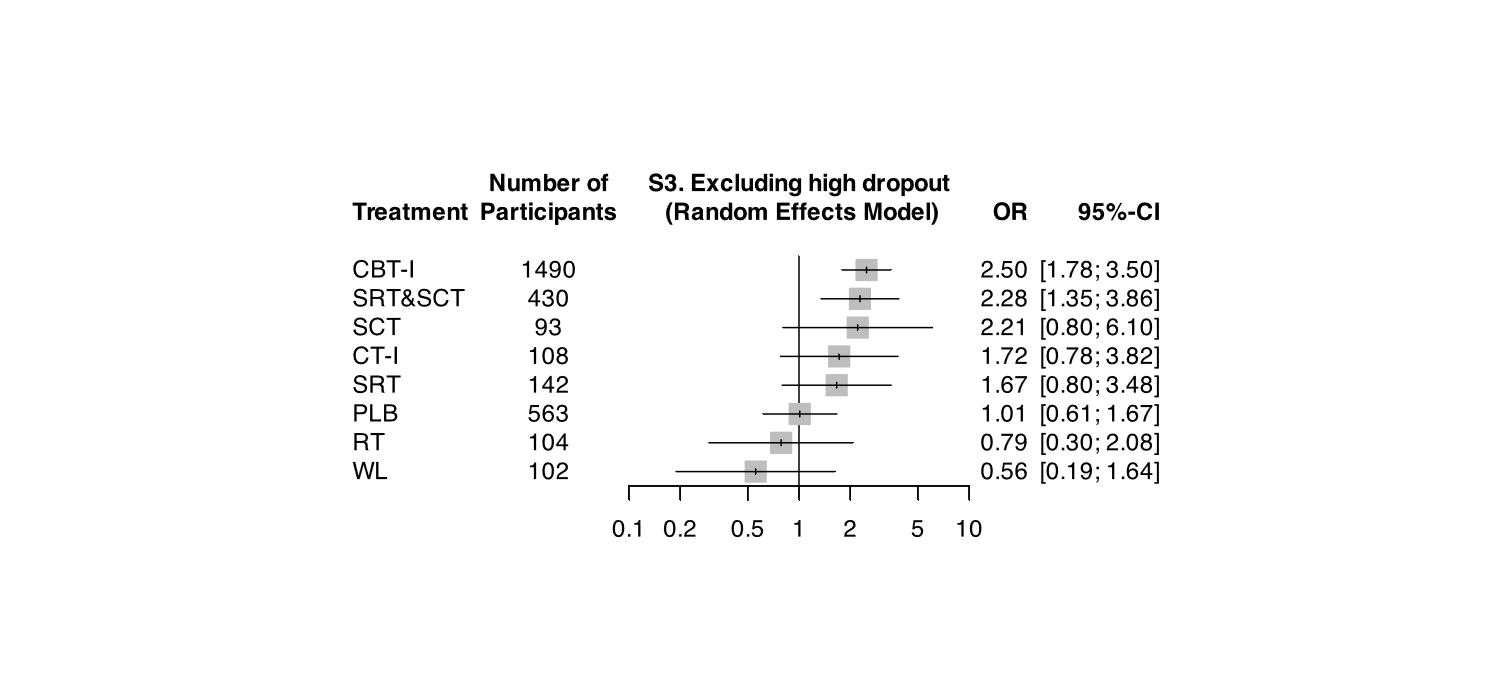

#### S4. To examine the influence of within-study bias

##### S4.1. Excluding trials with high RoB

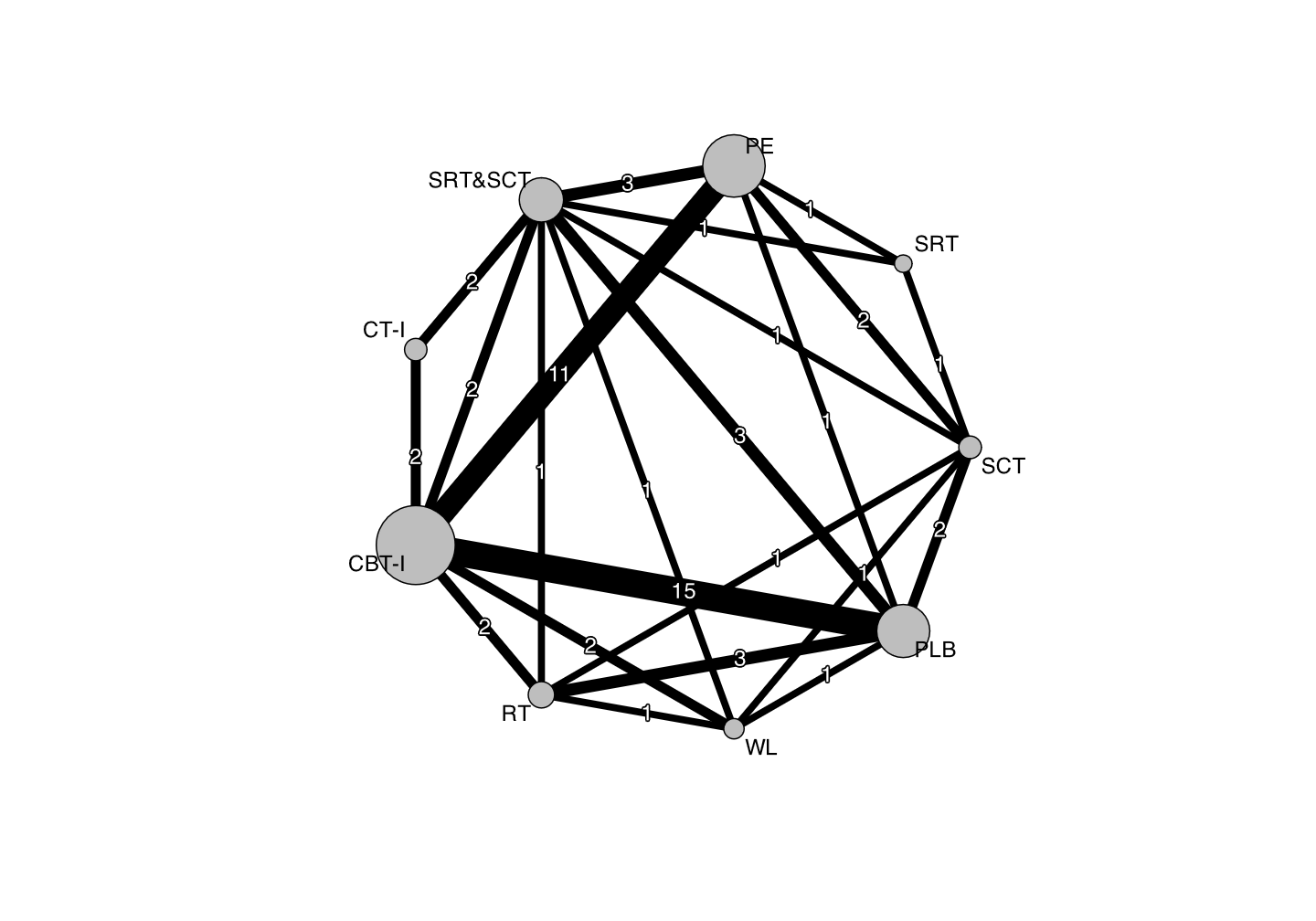

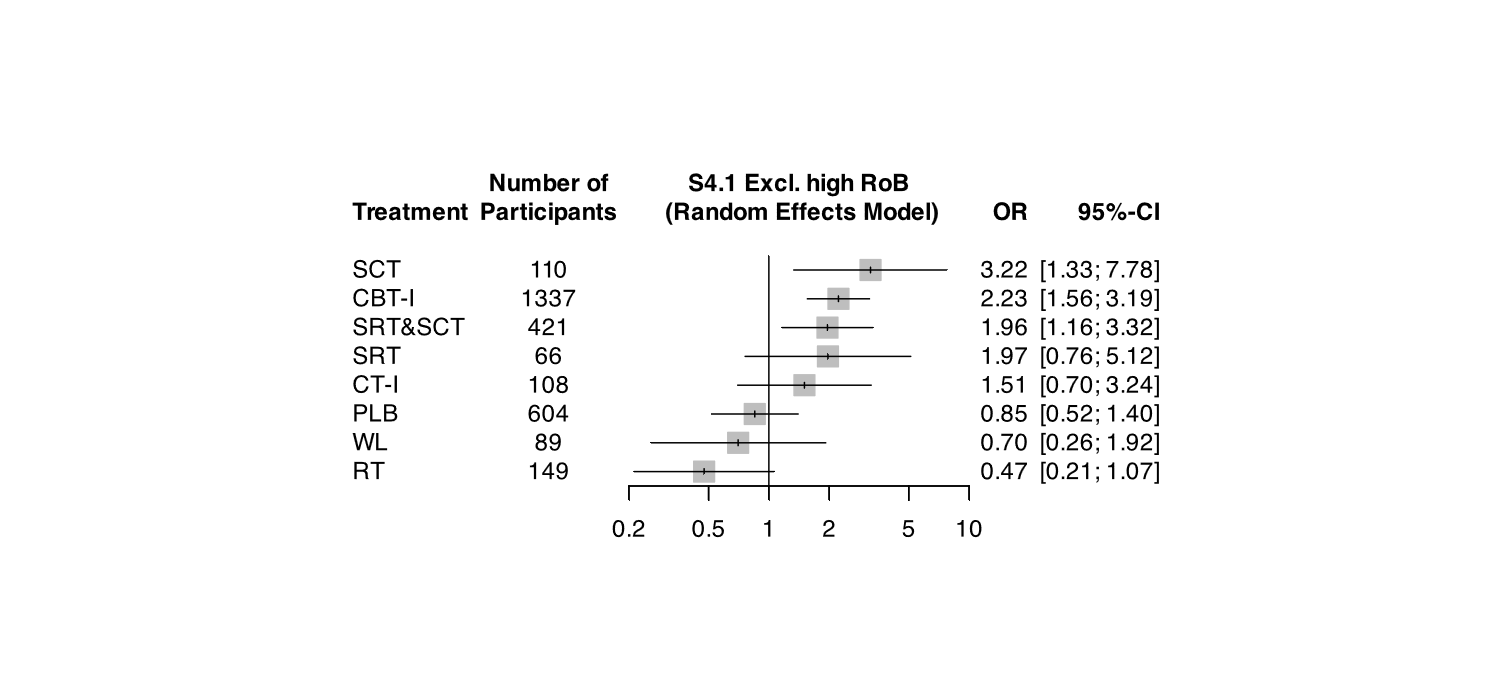

##### S4.2. Excluding trials with high and some concerns RoB

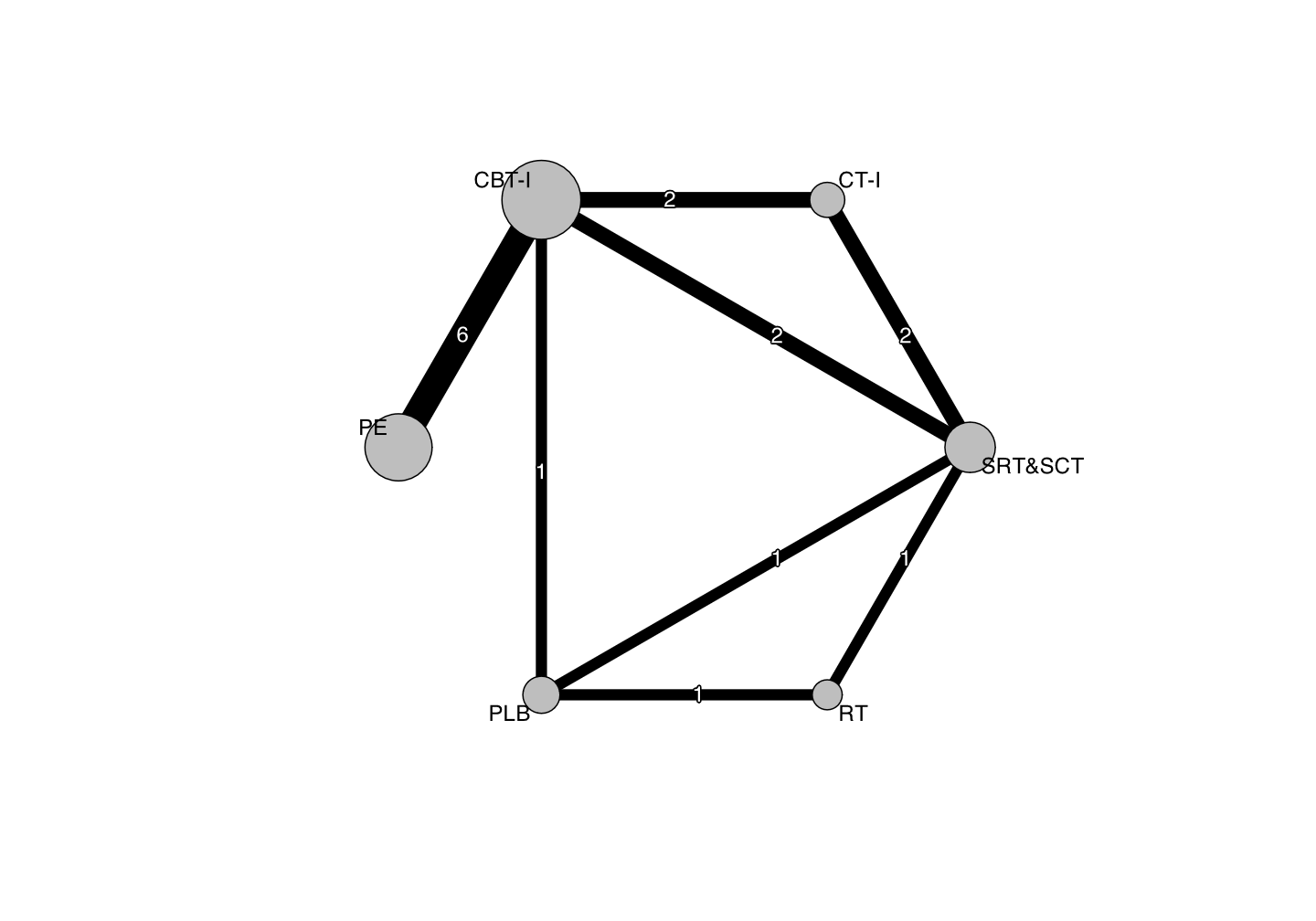

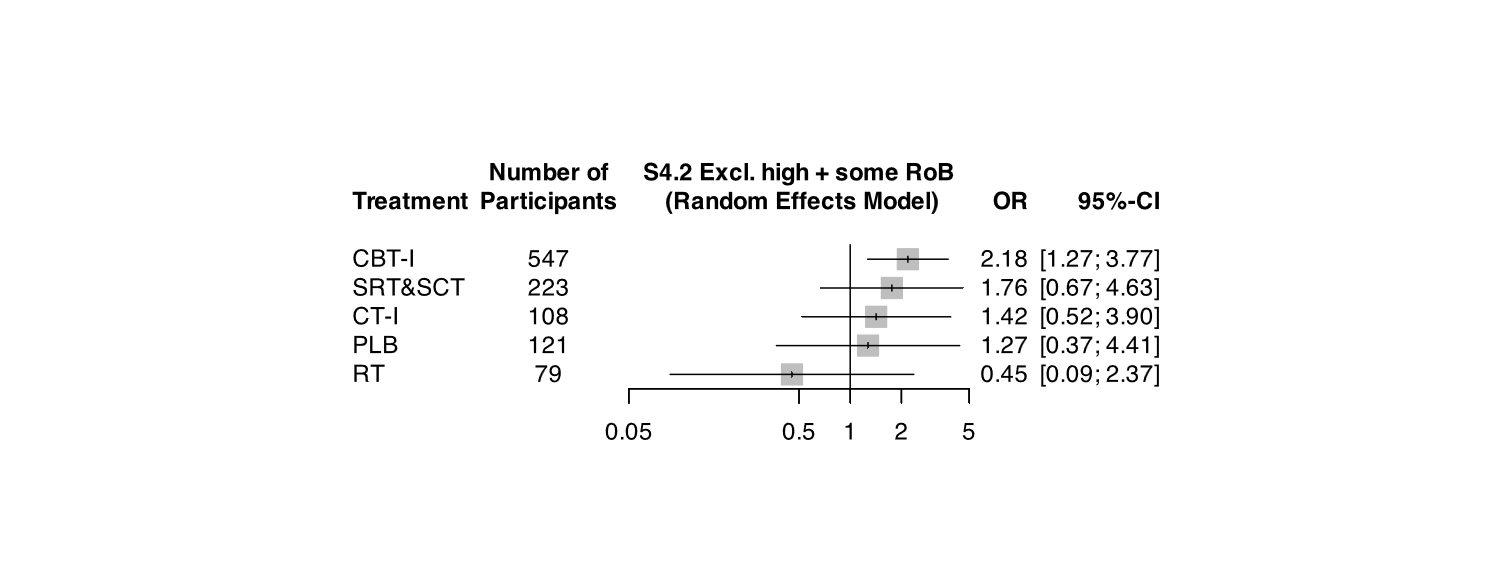

#### S5. Active arms only (vs RT)

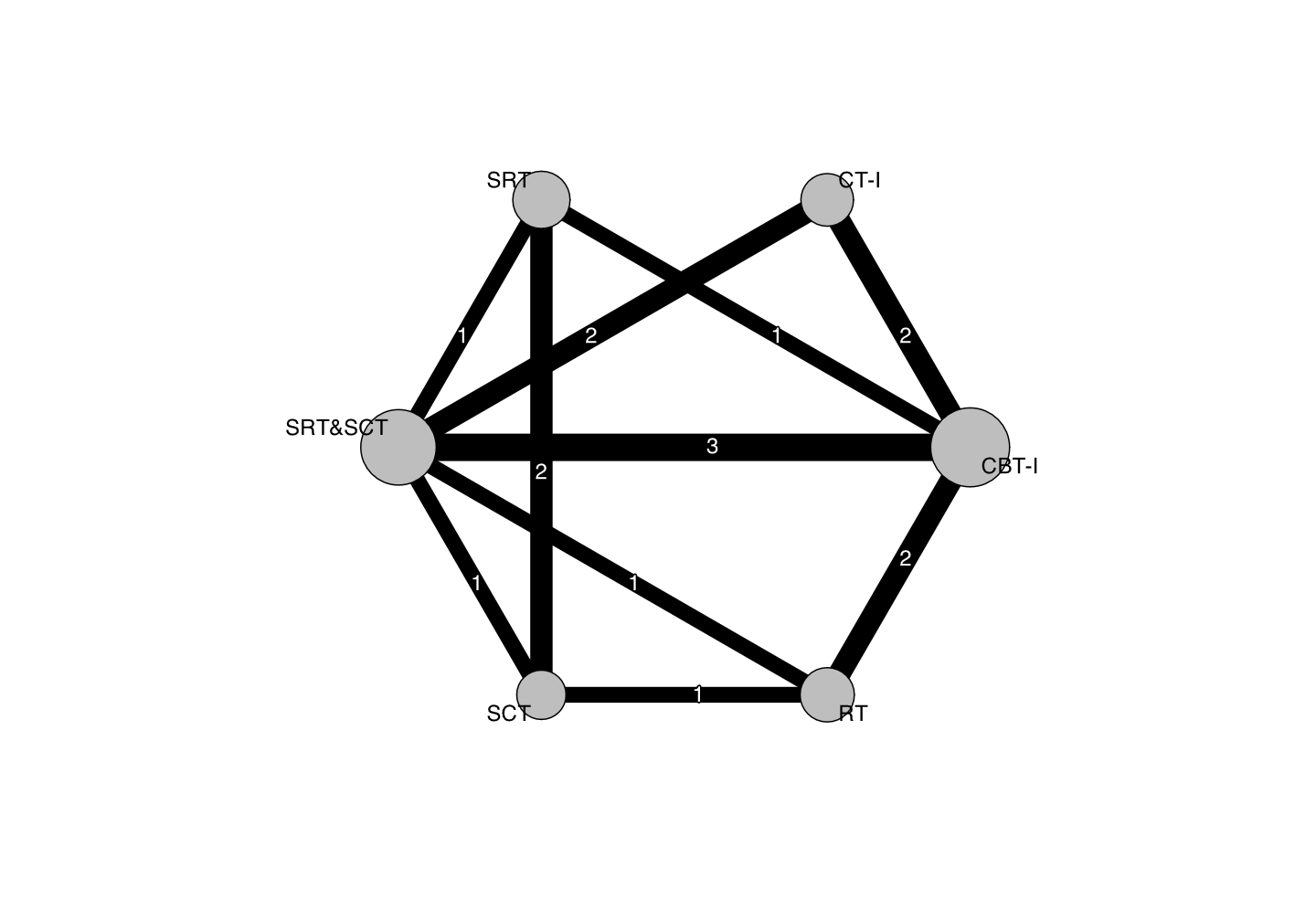

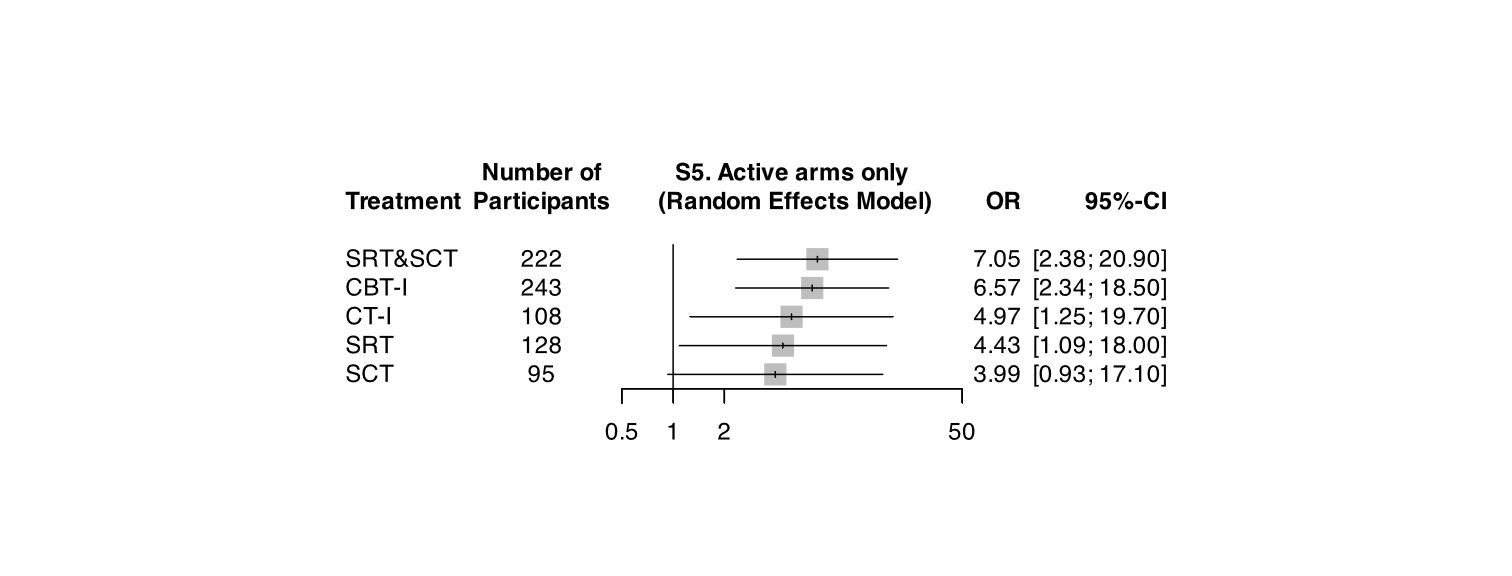

### 8. PRISMA-NMA

| **Section/Topic** | **Item #** | **Checklist Item** | **Reported on Page #** |
| --- | --- | --- | --- |
| **TITLE** |  |  |  |
| Title | 1 | Identify the report as a systematic review *incorporating a network meta-analysis (or related form of meta-analysis).* | TITLE PAGE |
| **ABSTRACT** |  |  |  |
| Structured summary | 2 | Provide a structured summary including, as applicable:  **Background:** main objectives  **Methods:** data sources; study eligibility criteria, participants, and interventions; study appraisal; and *synthesis methods, such as network meta-analysis.*  **Results:** number of studies and participants identified; summary estimates with corresponding confidence/credible intervals; *treatment rankings may also be discussed. Authors may choose to summarize pairwise comparisons against a chosen treatment included in their analyses for brevity.*  **Discussion/Conclusions:** limitations; conclusions and implications of findings.  **Other:** primary source of funding; systematic review registration number with registry name. | ABSTRACT |
| **INTRODUCTION** |  |  |  |
| Rationale | 3 | Describe the rationale for the review in the context of what is already known*, including mention of why a network meta-analysis has been conducted.* | MAIN TEXT / INTRODUCTION |
| Objectives | 4 | Provide an explicit statement of questions being addressed, with reference to participants, interventions, comparisons, outcomes, and study design (PICOS). | MAIN TEXT / INTRODUCTION |
| **METHODS** |  |  |  |
| Protocol and registration | 5 | Indicate whether a review protocol exists and if and where it can be accessed (e.g., Web address); and, if available, provide registration information, including registration number. | MAIN TEXT / METHODS  (https://osf.io/z48r2/) |
| Eligibility criteria | 6 | Specify study characteristics (e.g., PICOS, length of follow-up) and report characteristics (e.g., years considered, language, publication status) used as criteria for eligibility, giving rationale. *Clearly describe eligible treatments included in the treatment network, and note whether any have been clustered or merged into the same node (with justification).* | METHODS / Eligibility criteria |
| Information sources | 7 | Describe all information sources (e.g., databases with dates of coverage, contact with study authors to identify additional studies) in the search and date last searched. | METHODS / Search strategies |
| Search | 8 | Present full electronic search strategy for at least one database, including any limits used, such that it could be repeated. | SLEEPI; https://osf.io/c82xu/ |
| Study selection | 9 | State the process for selecting studies (i.e., screening, eligibility, included in systematic review, and, if applicable, included in the meta-analysis). | METHODS / Data extraction and risk of bias assessment, eAppendix2 |
| Data collection process | 10 | Describe method of data extraction from reports (e.g., piloted forms, independently, in duplicate) and any processes for obtaining and confirming data from investigators. | METHODS / Data extraction and risk of bias assessment |
| Data items | 11 | List and define all variables for which data were sought (e.g., PICOS, funding sources) and any assumptions and simplifications made. | SLEEPI; https://osf.io/c82xu/ |
| **Geometry of the network** | **S1** | Describe methods used to explore the geometry of the treatment network under study and potential biases related to it. This should include how the evidence base has been graphically summarized for presentation, and what characteristics were compiled and used to describe the evidence base to readers. | METHODS / Statistical analysis |
| Risk of bias within individual studies | 12 | Describe methods used for assessing risk of bias of individual studies (including specification of whether this was done at the study or outcome level), and how this information is to be used in any data synthesis. | METHODS / Data extraction and risk of bias assessment |
| Summary measures | 13 | State the principal summary measures (e.g., risk ratio, difference in means). *Also describe the use of additional summary measures assessed, such as treatment rankings and surface under the cumulative ranking curve (SUCRA) values, as well as modified approaches used to present summary findings from meta-analyses.* | METHODS / Data extraction and risk of bias assessment |
| Planned methods of analysis | 14 | Describe the methods of handling data and combining results of studies for each network meta-analysis. This should include, but not be limited to:   - *Handling of multi-arm trials;* - *Selection of variance structure;* - *Selection of prior distributions in Bayesian analyses; and* - *Assessment of model fit.* | METHODS / Statistical analysis |
| **Assessment of Inconsistency** | **S2** | Describe the statistical methods used to evaluate the agreement of direct and indirect evidence in the treatment network(s) studied. Describe efforts taken to address its presence when found. | METHODS / Statistical analysis |
| Risk of bias across studies | 15 | Specify any assessment of risk of bias that may affect the cumulative evidence (e.g., publication bias, selective reporting within studies). | METHODS / Statistical analysis |
| Additional analyses | 16 | Describe methods of additional analyses if done, indicating which were pre-specified. This may include, but not be limited to, the following:   - Sensitivity or subgroup analyses; - Meta-regression analyses; - *Alternative formulations of the treatment network; and* - *Use of alternative prior distributions for Bayesian analyses (if applicable).* | METHODS / Statistical analysis |
| **RESULTS†** |  |  |  |
| Study selection | 17 | Give numbers of studies screened, assessed for eligibility, and included in the review, with reasons for exclusions at each stage, ideally with a flow diagram. | RESULTS |
| **Presentation of network structure** | **S3** | Provide a network graph of the included studies to enable visualization of the geometry of the treatment network. | Figure 1 |
| **Summary of network geometry** | **S4** | Provide a brief overview of characteristics of the treatment network. This may include commentary on the abundance of trials and randomized patients for the different interventions and pairwise comparisons in the network, gaps of evidence in the treatment network, and potential biases reflected by the network structure. | RESULTS |
| Study characteristics | 18 | For each study, present characteristics for which data were extracted (e.g., study size, PICOS, follow-up period) and provide the citations. | RESULTS |
| Risk of bias within studies | 19 | Present data on risk of bias of each study and, if available, any outcome level assessment. | eAppendix2 |
| Results of individual studies | 20 | For all outcomes considered (benefits or harms), present, for each study: 1) simple summary data for each intervention group, and 2) effect estimates and confidence intervals. *Modified approaches may be needed to deal with information from larger networks.* | eAppendix5 |
| Synthesis of results | 21 | Present results of each meta-analysis done, including confidence/credible intervals. *In larger networks, authors may focus on comparisons versus a particular comparator (e.g. placebo or standard care), with full findings presented in an appendix. League tables and forest plots may be considered to summarize pairwise comparisons.* If additional summary measures were explored (such as treatment rankings), these should also be presented. | Figure3-5 |
| **Exploration for inconsistency** | **S5** | Describe results from investigations of inconsistency. This may include such information as measures of model fit to compare consistency and inconsistency models, *P* values from statistical tests, or summary of inconsistency estimates from different parts of the treatment network. | RESULTS, eAppendix3 |
| Risk of bias across studies | 22 | Present results of any assessment of risk of bias across studies for the evidence base being studied. | RESULTS, eAppendix2 |
| Results of additional analyses | 23 | Give results of additional analyses, if done (e.g., sensitivity or subgroup analyses, meta-regression analyses*, alternative network geometries studied, alternative choice of prior distributions for Bayesian analyses,* and so forth). | RESULTS, Figure5, eAppendix7 |
| **DISCUSSION** |  |  |  |
| Summary of evidence | 24 | Summarize the main findings, including the strength of evidence for each main outcome; consider their relevance to key groups (e.g., healthcare providers, users, and policy-makers). | DISCUSSION |
| Limitations | 25 | Discuss limitations at study and outcome level (e.g., risk of bias), and at review level (e.g., incomplete retrieval of identified research, reporting bias). *Comment on the validity of the assumptions, such as transitivity and consistency. Comment on any concerns regarding network geometry (e.g., avoidance of certain comparisons).* | DISCUSSION |
| Conclusions | 26 | Provide a general interpretation of the results in the context of other evidence, and implications for future research. | DISCUSSION |
| **FUNDING** |  |  |  |
| Funding | 27 | Describe sources of funding for the systematic review and other support (e.g., supply of data); role of funders for the systematic review. This should also include information regarding whether funding has been received from manufacturers of treatments in the network and/or whether some of the authors are content experts with professional conflicts of interest that could affect use of treatments in the network. | FUNDING |
